# Cognitive correlates of psychopathology in Functional/Dissociative Seizures and non-lesional epilepsy: an exploratory study

**DOI:** 10.1101/2024.04.24.24306276

**Authors:** Irene Faiman, Allan H. Young, Paul Shotbolt

## Abstract

**Objective:** To explore the relationship between cognitive functioning and psychopathological features in Functional/Dissociative Seizures (FDS), and test whether this differs from that observed in epilepsy.

**Methods:** We recruited a cross-sectional sample of adults (age > 18) with a diagnosis of non-lesional epilepsy or FDS between January 2021 and July 2022. Participants completed a series of psychiatric questionnaires and neuropsychological measures. Spearman’s Correlation Coefficient was computed between each of the psychiatric and cognitive measures in each group. Fisher’s Z test of significance for independent correlation coefficients then tested the significance of the difference between correlation coefficients for the two groups.

**Results:** There were no group differences in neuropsychological test scores. However, people with FDS reported higher seizure severity, depression levels, number of medically unexplained somatic symptoms, and exposure to traumatic events compared to epilepsy. Results of the Fisher’s Z-test revealed significant differences in correlation coefficients between groups in two instances. First, in the association between the number of traumatic experiences and cognitive switching (z = 2.77, p = 0.006); the number of traumatic experiences were positively associated with cognitive switching in epilepsy but showed a non-significant negative trend in FDS. Secondly, in the association between vocabulary abilities and the number of medically unexplained symptoms (z = -2.71; p = 0.007); higher vocabulary ability was associated with fewer somatic symptoms in epilepsy, while no such correlation was observed in FDS.

**Significance:** This study provides preliminary evidence for the complex interplay between cognitive functioning and psychopathology in FDS and epilepsy. Neurocognitive functioning such as vocabulary abilities or attentional switching may play a role in the expression or maintenance of pathological features of FDS.

**KEY POINTS:**

- People with non-lesional epilepsy or Functional/Dissociative Seizures (FDS) perform more poorly than healthy controls on neuropsychological measures.
- It is often thought that psychopathological factors influence cognitive presentation in FDS, but this hypothesis has received little empirical support.
- This study explores the relationship between cognition and psychopathology in FDS and epilepsy.
- Correlation analyses reveal distinct associations in FDS compared to epilepsy, suggesting potential differences in underlying mechanisms.
- Neurocognitive processes such as vocabulary abilities or attentional switching might contribute to FDS generation or presentation.

## INTRODUCTION

Cognitive functioning is supported by a complex and multi-dimensional network system. Characterising cognitive profiles in neurological and neuropsychiatric populations such as epilepsy and Functional/Dissociative Seizures (FDS(1)) is challenging as multiple interacting factors can affect cognition, resulting in a wide plethora of cognitive presentations.

Despite the importance of identifying and addressing cognitive impairment in epilepsy and FDS, this goes often undetected or undocumented (2). To date, there is still no consensus on an overall taxonomy of cognitive disorders in epilepsy, nor on a common battery of neuropsychological tests to use in diagnostic epilepsy services (3). Knowledge on the cognitive characteristics of FDS is even earlier in its infancy, as the existing literature concerning the neuropsychology of FDS was only recently systematically summarised, and described as particularly inconsistent and scarce (4). Small sample sizes avert the exploration of patient sub-groups and the identification of cognitive phenotypes, and failure to integrate psychiatric information in neuropsychological studies further limits our understanding.

The picture emerging from the evidence published so far indicates that both people with non-lesional epilepsy and people with FDS perform more poorly than healthy controls on a wide range of neuropsychological measures (5,6). There is also evidence of people with FDS outperforming people with epilepsy on measures of verbal comprehension (7,8), attention and working memory(8–10), performance validity (8,11) and phonemic fluency (8,10).

Evidence is more consistent in highlighting that people with FDS have significantly higher depression rates, complaints about physical symptoms, and reports of exposure to traumatic events than people with epilepsy, with some evidence also supporting a higher tendency to report dissociative experiences in daily life (12).

The reasons why people with FDS show similar impairment to people with epilepsy on cognitive measures in the absence of an “organic” disorder accounting for neuropsychological deficits are not clear. Different theories have been advanced such as a higher influence of emotional processes and psychopathology (13), or misdirected effort or attention (4,14).

However, despite theories advocating a psychiatric aetiology of cognitive impairment in FDS, this hypothesis has received little empirical support. Evidence from other clinical populations such as fibromyalgia or chronic fatigue syndrome do not always support a link between psychopathology and cognitive difficulties (15,16).

The few existing studies on this that we were able to identify have exclusively focused on the influence of mood and anxiety on cognitive functioning (17–20) but have never examined factors that are central to the psychopathology of FDS such as dissociation, somatisation, or exposure to traumatic events. It is therefore unclear to what extent psychiatric symptomatology contributes to cognitive functioning in FDS, and whether this goes above and beyond the impact it has in other clinical populations, such as people with epilepsy. In epilepsy, evidence on the impact of psychiatric disorders on cognition is also lacking, and this was identified as a field with insufficient evidence by a meta-analysis on cognition in Idiopathic Generalised Epilepsy (5).

The aim of this study is to characterise the relationship between cognition and psychopathology in FDS and epilepsy. We explore to what extent measures of depression, anxiety, dissociation, somatisation, suicidality, and exposure to traumatic events correlate with cognitive functioning in FDS as compared to epilepsy. This is an exploratory study that might serve as a basis for future hypothesis formulation and power calculations.

## MATERIALS AND METHODS

### Participants

A cross-sectional non-consecutive sample was recruited between January 2021 and July 2022 from South London and Maudsley Hospital’s Department of Neuropsychiatry (Outpatients), the Lishman Unit, King’s College Hospital’s Department of Neurology (Outpatients), the EEG Department or the Activation Clinic. All recruitment procedures were carried out remotely due to COVID-19 restrictions on in-person attendance to the clinics for non-essential workers.

We recruited adults (over 18 years) with suspected or diagnosed and active Functional/Dissociative Seizures or epilepsy. A diagnosis of epilepsy was a clinical diagnosis as per operational clinical definitions(21). A diagnosis of FDS was determined either through video-EEG supported diagnosis according to ILAE definitions(1), or in cases lacking video-EEG confirmation, by strong clinical indicators of FDS. For inclusion in the study under a diagnosis of FDS, seizure-like phenomena had to consist of paroxysmal, time-limited alterations in motor, sensory, autonomic, and/or cognitive functioning occurring in the context of a ‘collapsing’ presentation and/or impaired responsiveness. All patients were diagnosed following review by a consultant neurologist specialising in epilepsy and/or a consultant neuropsychiatrist specialising in functional neurological disorders.

Exclusion criteria were seizure disorder being secondary to another CNS disorder such as trauma or infection, evidence of MRI or CT abnormalities, relevant history of other neurological or vascular disorders, neurodevelopmental disorders, or brain surgery, concurrent severe mental health disorder such as major depressive disorder, psychosis, schizophrenia, or obsessive-compulsive disorder, and (suspected) concurrent epilepsy and FDS disorder.

The study was approved by the London Queen Square Research Ethics Committee (REC reference: 20/LO/0784, IRAS ID: 265164, latest amendment 28/09/2021). This study is exploratory in nature, so a sample size calculation was not performed.

### Study measures

Participants attended a single study visit remotely on Microsoft Teams. Cognitive performance was evaluated through a neuropsychological battery assessing the following cognitive domains: premorbid functioning (Test of Premorbid Functioning; TOPF (22)), general intellectual functioning (TOPF-estimated Wechsler Adult Intelligence Scale WAIS-IV Full-Scale Intelligence Quotient; FSIQ (23)), attention and working memory (WAIS-IV Digit Span and Arithmetic (24)), verbal comprehension and knowledge acquisition (WAIS-IV Vocabulary and Information (24)), speed of information processing (Symbol Digit Modality Test (SDMT) Oral version (25)), and executive functioning (Delis–Kaplan Executive Function System (D-KEFS) Verbal Fluency test (26)). The Letter – Category and Switching – Category Contrast scores were calculated (26). Performance validity was assessed by means of the Reliable Digit Span, with a cut-off score of 7.1 for effort test failure (27). Raw scores for these tests were demographically adjusted using test-specific normative tables, and then converted to Z scores. Participants had no known external incentives to fail on cognitive testing. We administered the SDMT vertically through a computer screen, deviating from guidelines; we acknowledge possible differences in perception and task response times due to the different format and reading orientation.

Questionnaires measuring the most defining psychopathological features of FDS were selected to explore their relationship with cognitive performance (12). These included measures of symptoms of depression (Patient Health Questionnaire-9; PHQ-9 (28)), anxiety (Generalised Anxiety Disorder Assessment-7; GAD-7 (29)), medically unexplained somatic symptoms (Screening for Somatoform Symptoms-2; SOMS-2 (30)), dissociation (Dissociative Experiences Scale; DES-II (31)), exposure to potentially traumatising events (Traumatic Experience Checklist; TEC (32)), and suicide risk (Mini International Neuropsychiatric Interview (MINI) Module C (33)). The Seizure Severity Questionnaire (SSQ) was used as an index of seizure severity over the past four weeks (34). Distress and satisfaction in relation to study participation were assessed on a 10-point Likert scale.

### Population description

Medication load for each patient was calculated for Antiepileptic Drugs (AEDs) and Antidepressants as the ratio of the prescribed daily dose to the defined daily dose (https://www.whocc.no/atc_ddd_index/), as determined by the WHO Collaborating Centre for Drug Statistics Methodology; when patients were taking multiple drugs of the same class, load scores were summed (35).

To estimate the significance of group differences for descriptive categorical variables, Chi-Squared test (36) was used, or Fisher’s exact test (37) in the presence of low count (n<5 in cross-tabulated cells). To estimate the significance of group differences for continuous variables, independent sample t-test was used in the presence of normal distribution, Mann-Whitney U test otherwise (38). Data were analysed using R statistical software version 4.0.0 (2020-04-24).

### Group comparison analyses

To estimate the significance of group differences in psychiatric and cognitive measures, the independent sample t-test was used in the presence of normal distribution, the Mann-Whitney U test otherwise (38). Results were compared against Bonferroni-corrected α = 0.007 for psychiatric questionnaires (0.05 divided by seven psychiatric questionnaires comparisons), and α = 0.004 for neuropsychological measures (0.05 divided by 14 neuropsychological measures).

### Correlation analyses

Separately for each group, the Spearman’s Correlation Coefficient was computed between each of the psychiatric and cognitive measures. The proportion of shared variance between variable’s ranks was calculated by squaring Spearman’s rho (ρ^2^) (39).

Analyses were performed following case-wise outlier exclusion (except for rare yet valid cases in skewed variables: TEC, MINI and SSQ). Potential outliers were defined as those points lying in the following interval, I = [q_0.25_ - 1.5 * IQR; q_0.75_ + 1.5 * IQR], where the IQR is the Interquartile Range, calculated as the difference between the 75^th^ (q_0.75_) and the 25^th^ quartiles (q_0.25_). In total, 13 individual values from 10 different variables were excluded. Analysis performed on the whole sample (including the 13 outlier values) are reported in Appendix S1 for comparison (Figure S1; Tables S8 to S13).

In order to test the significance of the difference between correlation coefficients for the two diagnostic groups, Spearman’s rho were transformed to Fisher’s z scores (40,41) using the R ‘cocor’ statistical package version 1.1-4 (2022-06-28; (42)). A Fisher’s Z test of significance for independent correlation coefficients was then run on converted scores (40,41). The Fisher Z test formula (40,41) was corrected to accommodate Spearman rank correlations instead of Pearson’s (43). The original expression 1/(n-3), as in the standard error of the Fisher Z difference, was replaced by 1.06/(n-3) as follows:

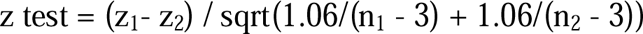

The denominator is the standard error of the difference (with n_1_ and n_2_ being the group sample sizes), and z_1_ and z_2_ are the z-transformed correlation coefficients for the two groups (r_1_ and r_2_), obtained based on the original formula:

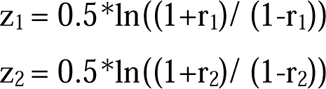

The number of Z tests performed between correlation coefficients was high (n = 84). Applying the Bonferroni correction would imply comparing p-values against a significance level α=0.0006. Given the study’s exploratory nature, the threshold for significance was relaxed to α=0.01. Results are discussed keeping into account the increased risk of Type I error.

To obtain an estimate of the effect size, Cohen’s q was calculated as the difference between two Fisher’s z-converted correlation coefficients; q < 0.1 represents no effect, 0.1 < q < 0.3 small, 0.3 < q < 0.5 medium, and q > 0.5 a large effect (44).

## RESULTS

### Population characteristics

Of the 81 people consenting, 8 were excluded due to failure to attend or uncertain diagnosis at time of analysis. 73 people were included in the analyses (epilepsy n = 34; FDS n = 39).

Gender, age, ethnicity, and marital status were comparable between groups, but people with FDS had significant fewer years of education, on average; consistently, a higher proportion of people with FDS reported suspected or diagnosed dyslexia (Table 1). Educational level was therefore controlled for in subsequent group comparison analyses. Substance use habits were similar, except for higher average alcohol consumption in the epilepsy group (Table 1).

**Table 1.**
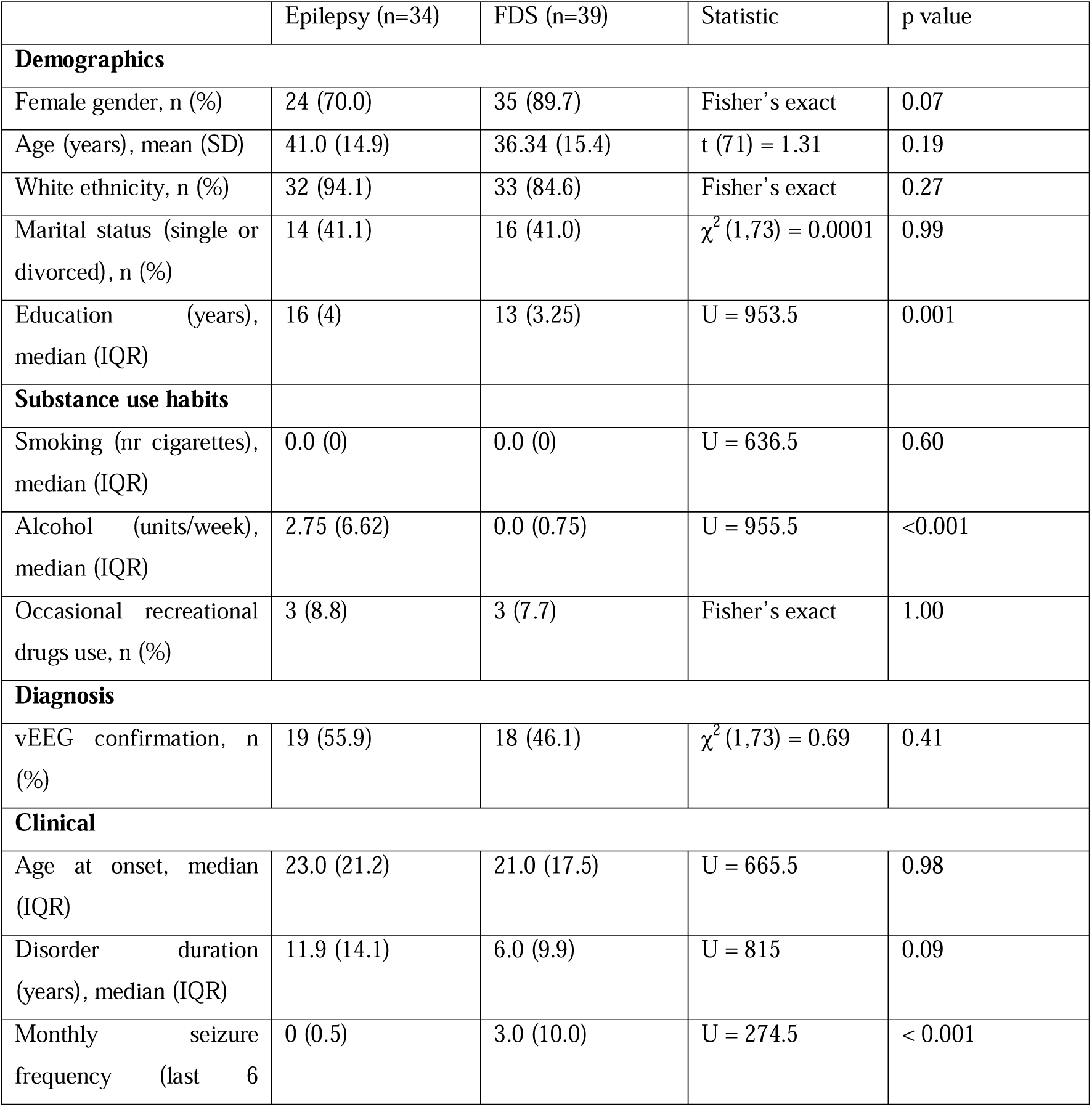
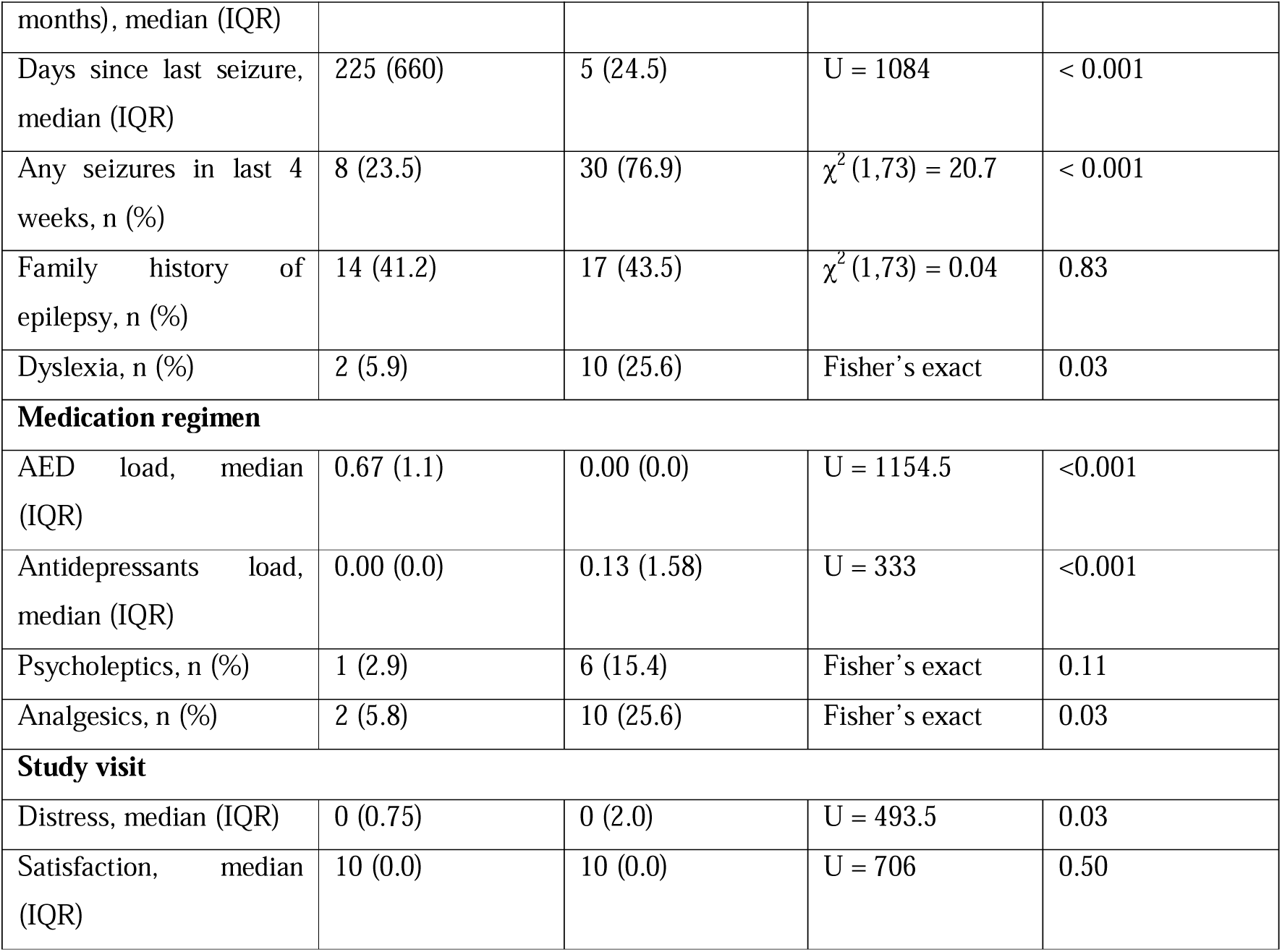
Demographic and clinical characteristics for 34 people with epilepsy and 39 people with FDS, with comparative statistics.

Approximately half of the sample in each group had video-EEG support for their diagnosis (Table 1). For FDS, this consisted of recording of a typical event in the absence of EEG changes (1); for epilepsy, this consisted of capturing interictal epileptiform abnormalities or seizures on EEG. All epilepsy patients had unknown seizure aetiology. 16 were classified as having probable focal epilepsy, 10 as probable generalised epilepsy; eight had unknown epilepsy type.

Age at seizure onset and disorder duration were comparable between groups. However, the FDS group reported significantly higher seizure frequency and lower latency from the last seizure than the epilepsy group, suggesting poorer seizure control in FDS (Table 1). Consistently, only a quarter of people with epilepsy reported experiencing seizures in the month prior to the study visit, which was significantly lower than rates reported by people with FDS. A comparable proportion of people reported a family history of epilepsy across groups (Table 1).

As would be expected, people with epilepsy had a significantly higher antiepileptic drug load intake, whilst people with FDS had a significantly higher antidepressant load intake. A higher proportion of people with FDS were taking analgesics. There was no difference in reported psycholeptics intake (including anxiolytics, antipsychotics, hypnotics and sedatives).

### Group comparison analyses

No group differences were observed in any of the neuropsychological test scores, after controlling for educational levels (Table 2). Uncontrolled comparisons are reported in Table S1. A higher number of people with FDS failed effort testing, but this was not significantly different from failure rates in epilepsy; Reliable Digit Span mean scores were not significantly different after Bonferroni correction (Table 2).

**Table 2.** Results of group comparison analysis for each variable. ANCOVA analyses on neuropsychological measures are adjusted for educational level. Asterisks indicate a significant difference in Bonferroni-corrected comparisons.

|  | Epilepsy (n=34) | FDS (n=39) | Statistic | p value |
| --- | --- | --- | --- | --- |
| **Seizure severity** |  |  |  |  |
| SSQ, median (IQR) | 0 (0.0) | 3.07 (2.8) | U = 240.5 | < 0.001* |
| **Psychiatric questionnaires** |  |  |  |  |
| GAD-7, median (IQR) | 5.5 (7.0) | 8.0 (7.5) | U = 465 | 0.03 |
| PHQ-9, median (IQR) | 5.0 (6.0) | 10.0 (9.0) | U = 324.5 | < 0.001* |
| MINI, median (IQR) | 0.0 (0.0) | 1.0 (4.0) | U = 474.5 | 0.02 |
| SOMS-II, median (IQR) | 5.0 (8.5) | 17.0 (10.5) | U = 288 | < 0.001* |
| DES-2, median (IQR) | 5.71 (12.0) | 15.0 (18.75) | U = 435.5 | 0.01 |
| TEC, median (IQR) | 2.0 (3.75) | 5.0 (4.5) | U = 320 | < 0.001* |
| TEC pre-symptoms onset, median (IQR) | 1.0 (3.0) | 5.0 (3.5) | U = 284 | < 0.001* |
| **Neuropsychological measures** |  |  |  |  |
| Reliable Digit Span, mean (SD) | 9.88 (2.32) | 8.54 (1.73) | F (1,70) = 4.16 | 0.04 |
| Effort test failure (RDS < 7.1), n (%) | 4 (11.7) | 11 (28.2) | Fisher's exact | 0.14 |
| TOPF-derived FSIQ, z mean (SD) | 0.02 (0.57) | -0.34 (0.63) | F (1,70) = 2.25 | 0.14 |
| Digit Span, z mean (SD) | -0.01 (0.95) | -0.48 (0.79) | F (1,70) = 2.97 | 0.09 |
| Arithmetic, z mean (SD) | 0.03 (1.07) | -0.42 (0.97) | F (1,70) = 0.50 | 0.48 |
| Working Memory Index, z mean (SD) | 0.01 (1.00) | -0.54 (0.93) | F (1,70) = 2.13 | 0.15 |
| Vocabulary, z mean (SD) | 0.46 (0.96) | 0.19 (1.20) | F (1,70) = 0.05 | 0.83 |
| Information, z mean (SD) | 0.38 (0.87) | 0.13 (1.28) | F (1,70) = 0.002 | 0.96 |
| Verbal Comprehension Index, z mean (SD) | 0.47 (0.98) | 0.19 (1.33) | F (1,70) = 0.03 | 0.85 |
| SDMT, z mean (SD) | -0.90 (1.29) | -0.81 (1.31) | F (1,70) = 0.53 | 0.47 |
| Letter Fluency, z mean (SD) | 0.16 (1.35) | -0.25 (1.08) | F (1,70) = 1.20 | 0.28 |
| Category Fluency, z mean (SD) | 0.68 (1.36) | 0.42 (1.17) | F (1,70) = 0.13 | 0.72 |
| Category Switching, z mean (SD) | 0.55 (1.19) | 0.47 (1.36) | F (1,70) = 0.17 | 0.68 |
| Contrast Letter - Category, z mean (SD) | -0.53 (1.02) | -0.68 (1.15) | F (1,70) = 0.75 | 0.39 |
| Contrast Switching - Category, z mean (SD) | -0.09 (1.22) | 0.07 (1.09) | F (1,70) = 0.60 | 0.44 |

Compared to age-matched normative data, mean z scores for both groups fell within the average range [-0.67, 0.67] for all neuropsychological measures except processing speed (SDMT). Scores on this measure were expected to be lower due to a deviation from administration guidelines to allow remote assessment mode (see methods).

People with FDS reported significantly higher seizure severity over the past 4 weeks than people with epilepsy (Table 2). Compared against Bonferroni-corrected α = 0.007, people with FDS reported significantly higher depression levels, medically unexplained somatic complains, and number of potentially traumatising events, including when only considering events occurring before symptom onset. Levels of anxiety, dissociative experiences, and suicide risk were significantly higher in FDS in Bonferroni-uncorrected comparisons only (Table S1).

### Correlation analyses

As a first step, Spearman’s correlations between psychiatric and neuropsychological measures were computed separately for each group. Results that were significant against α = 0.01 are reported descriptively. Results for the remainder of the correlations tested are reported graphically in Figure 1; further details (Correlation Coefficients and p-values) are in Tables S2 - S7.

**Figure 1.**
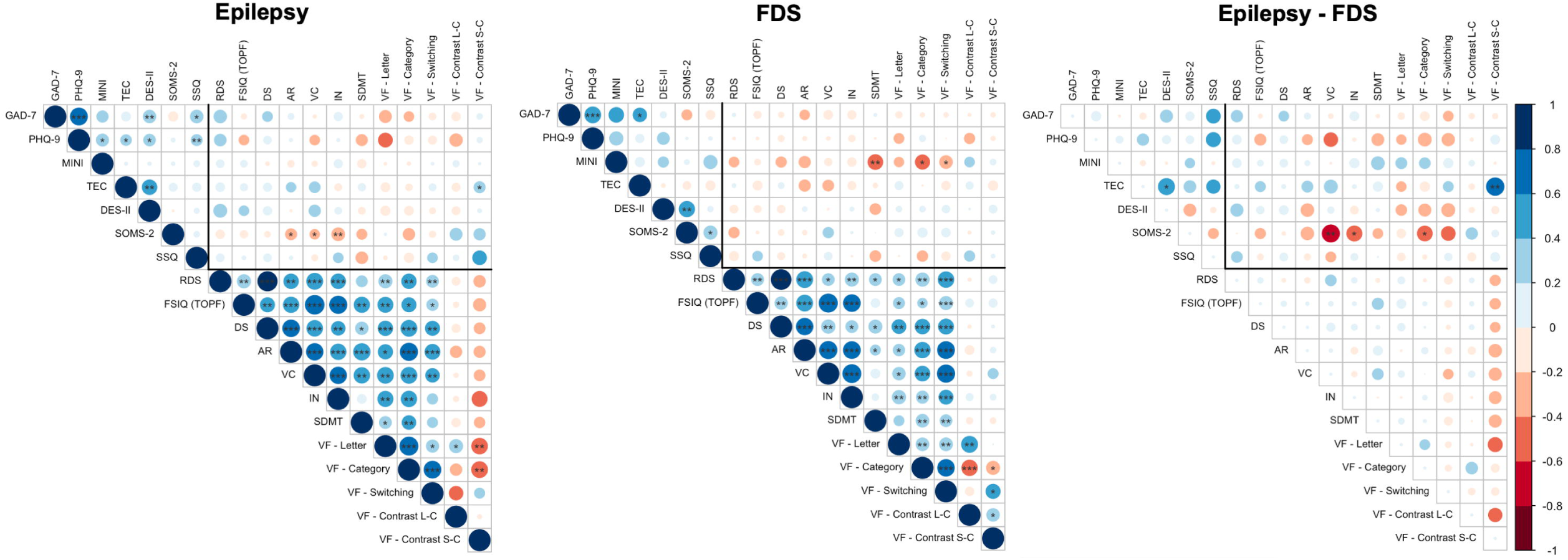
Correlation matrices for people with epilepsy (left), people with FDS (middle) and for the difference in correlation coefficients between epilepsy and FDS (right), with associated significance levels (asterisks). Correlations of primary interest for this study are those within the black frame. * Significant at α = 0.05; ** Significant at α = 0.01; *** Significant at α = 0.001. Blue = positive association; Red = negative association; the colour intensity reflects the magnitude of the correlation coefficient (or the magnitude of the difference for the right matrix); Abbreviations (if not provided in text): RDS = Reliable Digit Span; DS = WAIS-IV Digit Span; AR = WAIS-IV Arithmetic; VC = WAIS-IV Vocabulary; IN = WAIS-IV Information; VF = Verbal Fluency; L-C = Letter – Category Contrast score; S-C = Switching – Category Contrast score.

In the group with epilepsy, scores on the WAIS-IV Information subtest were negatively correlated with the number of medically unexplained symptoms, as measured by the SOMS-2 (ρ(31) = -0.48; p = 0.004; Figure 1). When considering correlations between psychiatric measures in epilepsy, seizure severity (SSQ) was positively associated with depression (PHQ-9) scores (ρ(31) = 0.44; p = 0.01). Dissociative symptoms (DES-II) significantly correlated with both anxiety (GAD-7; ρ(30) = 0.46; p = 0.009) and number of traumatic experiences (TEC; ρ(30) = 0.54; p = 0.001). GAD-7 and PHQ-9 scores were highly correlated (ρ(31) = 0.71; p < 0.001). The proportion of shared variance (ρ^2^) between the ranks of these significantly correlated variables ranged between 19% and 50% (Table S4).

In the group with FDS, suicide risk scores (MINI) were significantly and negatively correlated with processing speed scores (SDMT; ρ(36) = -0.44; p = 0.005; Figure 1). When examining correlations between psychiatric measures in FDS, dissociative symptoms (DES-II) were significantly and positively correlated with the number of medically unexplained symptoms (SOMS-2; ρ(36) = 0.48; p = 0.002). GAD-7 and PHQ-9 scores were positively correlated (ρ(37) = 0.51; p = 0.001). The proportion of shared variance (ρ^2^) ranged between 19% and 26% (Table S2).

Scores of several neuropsychological tests were highly intercorrelated in both groups (Figure 1, Tables S2 – S5).

Results of the Fisher’s Z-test for the difference between correlation coefficients indicated that z-transformed rho values significantly differed between groups in two instances (Figure 1, Figure 2). First, there was a significant difference between groups in the association between the number of traumatic experiences (TEC) and a measure of cognitive switching (D-KEFS Verbal Fluency Switching - Category Contrast score; z = 2.77, p = 0.006; Figure 2a). For people with epilepsy, a higher number of traumatic experiences was associated with better switching (ρ(32) = 0.37; p = 0.03; ρ^2^ = 14%); in FDS there was a nonsignificant trend towards a negative association (ρ(37) = -0.31; p = 0.05). The effect size for the difference between correlations was large (Cohen’s q = 0.68; CI: 0.2 – 1.06).

**Figure 2.**
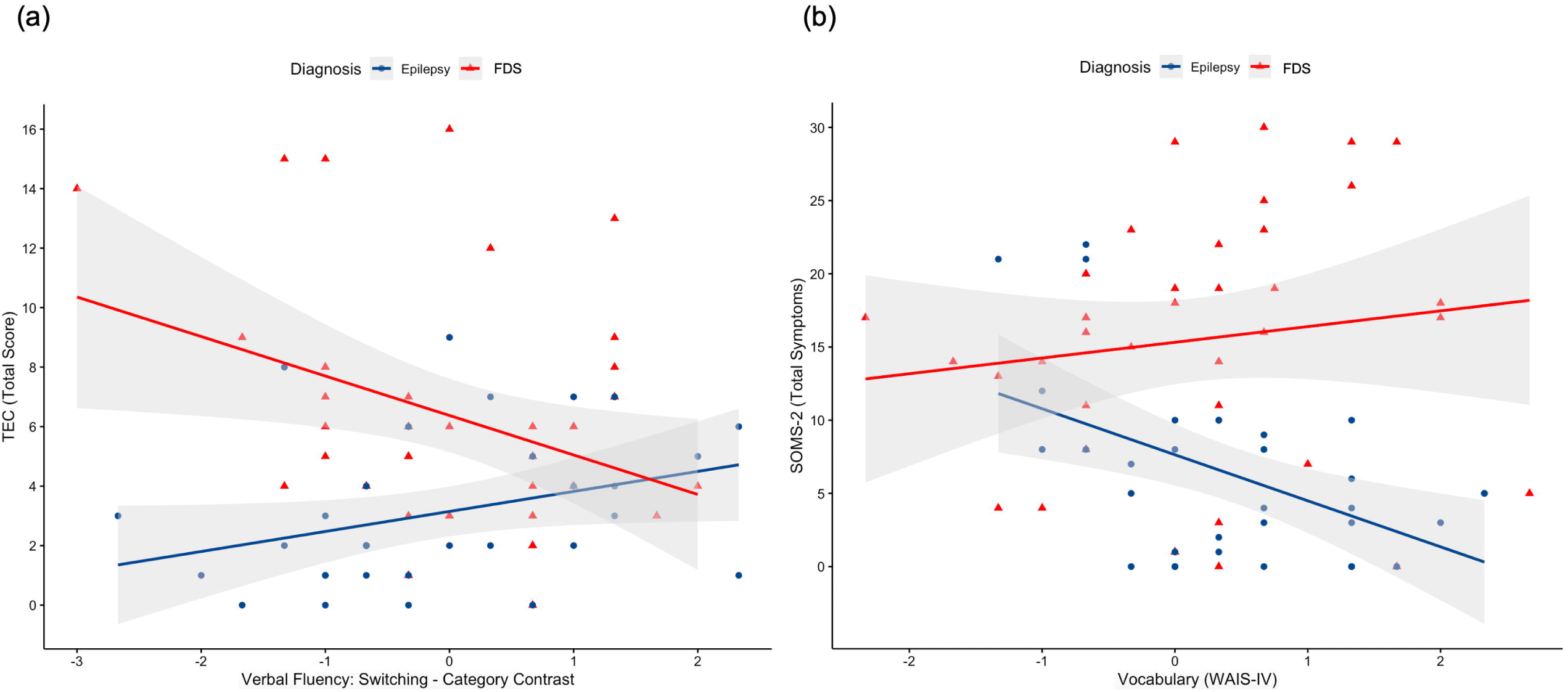
Scatterplots for the significant correlation differences at α = 0.01. (a) Group-wise correlation between the TEC and the Switching-Category Contrast score of the D-KEFS verbal fluency test (z score). (b) Group-wise correlation between the SOMS-2 questionnaire and the WAIS-IV Vocabulary test (z score). Linear regression models are fitted for visualisation purposes only. Variables on the x axis are z scores. TEC = Traumatic Experiences Checklist; SOMS-2 = Screening for Somatoform Symptoms-2.

Secondly, there was a significant difference between groups in the association between the WAIS-IV Vocabulary score and the number of medically unexplained symptoms (SOMS-2; z = -2.71; p = 0.007; Figure 2b). In epilepsy, as vocabulary abilities increased, there was a significant decrease in the number of unexplained somatic symptoms (ρ(31) = -0.41; p = 0.017; ρ^2^ = 17%). In FDS, no such correlation was observed (ρ(35) = 0.25; p = 0.14). The effect size for the difference between correlations was large (Cohen’s q = 0.66; CI: 0.18 – 1.05). Results for nonsignificant comparisons are in Tables S6 - S7.

## DISCUSSION

It is often postulated that neuropsychological performance in FDS is influenced by psychopathological processes; however, evidence in support of these claims is limited. This study explored the relationship between neuropsychological and psychiatric measures in FDS and assessed whether correlation patterns differ from those observed in a cohort of people with non-lesional epilepsy. Results suggest that the patterns of association between cognitive and psychiatric measures might provide useful information on the underlying disorders.

### Traumatic experiences and switching abilities

A significant group difference with large effect size was observed in the relationship between traumatic experiences and switching abilities. In people with epilepsy, a higher number of traumatic events was associated with greater switching ability, whilst the opposite trend was observed in FDS.

Attention switching is the ability to flexibly shift between mental sets and is essential for executive control. It has been consistently emphasised that people experiencing long-term consequences of trauma (e.g., PTSD or some people with FND), present with attentional bias towards threat-relevant stimuli, and difficulties disengaging from them (45).

Consistently, we observe that switching abilities in FDS have a different relationship with trauma than what is observed in epilepsy. A possibility is that cognitive differences might arise as a result of trauma; in people with epilepsy, a greater ability to disengage from a mental set might reflect the implementation of functional coping strategies. On the other hand, the ability of people with FDS to implement functional cognitive coping strategies might become reduced as a result of trauma, potentially facilitating the expression and maintenance of the disorder.

Another possibility is that this difference might reflect the existence of pre-trauma cognitive characteristics predisposing people to the subsequent development of FDS. For example, the experience of trauma in those with weaker switching abilities might contribute to difficulties in disengaging from traumatic stimuli, therefore increasing the chances of developing FDS. Overall, this finding provides some further support for the idea that weaknesses in attentional switching occur in interplay with traumatic experiences in FDS.

Although the effect size for the group difference was large, the amount of shared variance between the TEC score and the switching contrast score was relatively low in both groups (epilepsy: 14%; FDS: 10%), suggesting that other unaccounted factors such as subjective impact of trauma or emotional regulation strategies might play a relevant role.

### Vocabulary abilities and medically unexplained symptoms

A significant group difference with large effect size was found in the relationship between the number of medically unexplained physical symptoms and vocabulary abilities. In epilepsy, better vocabulary and verbal expression abilities were associated with fewer medically unexplained somatic symptoms; no such association was observed in FDS.

Medically unexplained symptoms are often considered a measure of somatisation, assuming psychological origin when a medical condition cannot be identified. Somatisation or conversion symptoms are believed to serve the purpose of communicating distress, under the assumption that emotions, rather than being expressed symbolically through words, are communicated through physical symptoms (46). Based on this conceptualisation, a greater ability to express and articulate verbal information would result in reduced expression of distress through somatisation, as supported by alexithymia studies (47). Our finding that people with epilepsy experience significantly more somatic complains at lower vocabulary abilities corroborates these findings.

In contrast, we observed no such association in FDS, possibly due to sub-group variations. While some individuals may follow the epilepsy pattern, others might not, resulting in a null association at the group level. Another possibility is that, at least in an FDS sub-group, high somatisation tendences might persist despite proficient verbal expression.

People with epilepsy and people with FDS differ in the way they verbally convey somatic experiences, such as seizures (48). Although underlying mechanisms remain unclear, psychological predispositions or coping styles might also play a role, suggesting a complex interplay between verbal expression abilities and physical symptoms reporting.

Despite the large effect size, the amount of shared variance between vocabulary and somatic symptoms was relatively low in both groups (epilepsy: 17%; FDS: 6%), indicating that other factors such as personality, beliefs, or internal symbolic representations could be at play.

Additionally, unexplained physical symptoms could be attributable to hypervigilance or to an impaired ability to ignore bodily stimuli, rather than somatisation tendencies (12). However, the somatisation hypothesis is in part supported by a recent meta-analysis highlighting that people with FDS experience depression-related physical symptoms more than depression-related cognitive and emotional symptoms; this suggests an increased tendency to express distress in the form of somatic symptoms (49).

### Strengths and limitations

These findings carry a risk of Type I error due to the extensive number of comparisons tested, requiring further validation. Similarly, the lack of significance for most comparisons should also consider the increased probability of Type II error; null findings may result from either the absence of true differences or insufficient power to detect them.

Study strengths include good seizure control in most of the epilepsy sample, reducing bias due to frequent seizure activity on cognition.

Conducting assessment remotely may introduce some bias (e.g., due to different assessment environments, sources of domestic distractions or facilitative cues), but the literature suggests minimal impact for verbally administered tests; measures including visual material (e.g., SDMT) pose more challenges (50).

People who failed performance validity testing (PVT; epilepsy n = 4, FDS n = 11) were not excluded from the analysis as one objective was to study the association between PVT and other variables. A failed PVT implies that neuropsychological scores for a minority of the sample cannot be assumed to reflect true cognitive abilities but are only an indication of patients’ minimum capacities.

The associations between variables could be mediated or confounded by third variables, and this was not accounted for in the current study, which was meant to provide an exploratory account. It is also possible that multiple sub-groups exist within each cohort, and this might limit the detection of a group-level effect. Attention to qualitative features such as subjective cognitive performance might also contribute to an improved understanding (4,14).

## CONCLUSION

We provide preliminary evidence for the interplay between cognitive functioning and two relevant clinical characteristics of FDS, which differ from an epilepsy control group: the tendency to experience a high number of medically unexplained physical symptoms and the exposure to potentially traumatising events.

We suggest that it might be useful to consider cognitive functioning not only as passively influenced by psychological factors; neurocognitive processes such as vocabulary abilities or attentional switching might contribute to generating or maintaining different pathological features of FDS.

Given the novelty of the research question, we adopted an exploratory approach in the service of future hypotheses generation. Should these findings be replicated, they could help identifying a useful set of neuropsychological tests for the assessment of people with FDS as to date, there is no consensus on a clinical battery, although some proposals have been advanced (13). Interventions targeting neurocognitive processes such as mindfulness or attentional and language retraining might then be explored.

## ACKNOWLEDGEMENTS

This work was supported by the Bergqvist Charitable Trust through the Psychiatry Research Trust as a PhD scholarship to Irene Faiman. Professor Young’s independent research is funded by the National Institute for Health Research (NIHR) Biomedical Research Centre at South London and Maudsley NHS Foundation Trust and King’s College London.

## AUTHORS’ CONTRIBUTION

Irene Faiman: Conceptualization (lead); investigation (lead); methodology (lead); data curation (lead); formal analysis (lead); software (lead); project administration (lead); visualization (lead); writing - original draft preparation (lead). Allan H Young: Conceptualization (supporting); methodology (supporting); project administration (supporting); funding acquisition (supporting); resources (equal); supervision (equal); writing – review & editing (equal). Paul Shotbolt: Conceptualization (supporting); methodology (supporting); project administration (supporting); funding acquisition (lead); resources (equal); supervision (equal); writing – review & editing (equal).

## DATA AVAILABILITY

Anonymised data are available upon request for collaborative purposes.

## ETHICS APPROVAL STATEMENT

The study was approved by the London Queen Square Research Ethics Committee (REC: 20/LO/ 0784, IRAS ID: 265164, 28/09/2021).

## PATIENT CONSENT STATEMENT

All participants gave written informed consent for the study.

