## Appendix S1 for "Cognitive correlates of psychopathology in Functional/Dissociative Seizures and non-lesional epilepsy: an exploratory study"

This appendix contains:

- Results of independent samples t-tests on cognitive measures when **not** controlling for education levels (Table S1)
- Detailed results for the correlation analyses presented in the main manuscript (Tables S2 – S7, Figure S1),
- Derailed results for the correlation analyses including outliers (Tables S8 - S13).

**Table S1.** Results of independent samples t-tests on cognitive measures, not controlling for education levels.

|  | **Epilepsy (n=34)** | **FDS (n=39)** | **Statistic** | **p value** |
| --- | --- | --- | --- | --- |
| **Neuropsychological measures** | | | | |
| Reliable Digit Span, mean (SD) | 9.88 (2.32) | 8.54 (1.73) | t (71) = 2.83 | 0.006 |
| TOPF-derived FSIQ, z mean (SD) | 0.02 (0.57) | -0.34 (0.63) | t (71) = 2.53 | 0.01 |
| Digit Span, z mean (SD) | -0.01 (0.95) | -0.48 (0.79) | t (71) = 2.30 | 0.02 |
| Arithmetic, z mean (SD) | 0.03 (1.07) | -0.42 (0.97) | t (71) = 1.87 | 0.06 |
| Working Memory Index, z mean (SD) | 0.01 (1.00) | -0.54 (0.93) | t (71) = 2.41 | 0.02 |
| Vocabulary, z mean (SD) | 0.46 (0.96) | 0.19 (1.20) | t (71) = 1.05 | 0.29 |
| Information, z mean (SD) | 0.38 (0.87) | 0.13 (1.28) | t (71) = 0.96 | 0.34 |
| Verbal Comprehension Index, z mean (SD) | 0.47 (0.98) | 0.19 (1.33) | t (71) = 1.03 | 0.31 |
| SDMT, z mean (SD) | -0.90 (1.29) | -0.81 (1.31) | t (71) = -0.30 | 0.76 |
| Letter Fluency, z mean (SD) | 0.16 (1.35) | -0.25 (1.08) | t (71) = 1.42 | 0.16 |
| Category Fluency, z mean (SD) | 0.68 (1.36) | 0.42 (1.17) | t (71) = 0.87 | 0.38 |
| Category Switching, z mean (SD) | 0.55 (1.19) | 0.47 (1.36) | t (71) = 0.26 | 0.79 |
| Contrast Letter – Category, z mean (SD) | -0.53 (1.02) | -0.68 (1.15) | t (71) = 0.60 | 0.54 |
| Contrast Switching - Category, z mean (SD) | -0.09 (1.22) | 0.07 (1.09) | t (71) = -0.58 | 0.56 |

**Table S2**. Detailed results for correlation analysis when outliers are excluded from the data. Presented are the Spearman Correlation Coefficients for the cohort of people with FDS.

|  | **gad7_total** | **phq9_total** | **mini_total_risk_score** | **tec_total_presence_score** | **des2_average** | **soms2_symptoms_total_1to53** | **ssq_total_score_4weeksCorrected** | **Reliable_Digit_Span** | **topf_fsiq_z** | **ds_z** | **ar_z** | **vc_z** | **in_z** | **sdmt_z** | **vf_letter_z** | **vf_category_z** | **vf_switch_tot_z** | **vf_contrast_lc_z** | **vf_contrast_sc_z** |
| --- | --- | --- | --- | --- | --- | --- | --- | --- | --- | --- | --- | --- | --- | --- | --- | --- | --- | --- | --- |
| **gad7_total** | 1 | 0.51 | 0.31 | 0.32 | 0.13 | -0.11 | -0.04 | -0.07 | -0.11 | -0.05 | -0.19 | -0.11 | -0.15 | 0.01 | -0.07 | -0.03 | 0.01 | -0.05 | 0.04 |
| **phq9_total** | 0.51 | 1 | 0.29 | 0.08 | 0.26 | 0.08 | 0 | 0.02 | 0.07 | -0.05 | 0.02 | 0.11 | -0.02 | 0.06 | -0.12 | 0.15 | 0.11 | -0.23 | -0.04 |
| **mini_total_risk_score** | 0.31 | 0.29 | 1 | 0.26 | 0.19 | -0.03 | 0.26 | -0.11 | 0 | -0.16 | -0.28 | -0.14 | -0.14 | -0.44 | -0.22 | -0.37 | -0.34 | 0.15 | -0.02 |
| **tec_total_presence_score** | 0.32 | 0.08 | 0.26 | 1 | -0.04 | -0.15 | -0.18 | 0.1 | -0.08 | 0.1 | -0.29 | -0.24 | -0.09 | -0.12 | 0.13 | 0.05 | -0.14 | 0.05 | -0.31 |
| **des2_average** | 0.13 | 0.26 | 0.19 | -0.04 | 1 | 0.48 | 0.14 | -0.09 | -0.06 | -0.07 | 0.14 | -0.01 | 0.13 | -0.14 | 0.15 | 0.14 | 0.2 | 0 | 0.17 |
| **soms2_symptoms_total_1to53** | -0.11 | 0.08 | -0.03 | -0.15 | 0.48 | 1 | 0.41 | -0.25 | -0.05 | -0.09 | -0.05 | 0.25 | 0.02 | -0.01 | 0.18 | 0.18 | 0.17 | -0.01 | 0.01 |
| **ssq_total_score_4weeksCorrected** | -0.04 | 0 | 0.26 | -0.18 | 0.14 | 0.41 | 1 | -0.23 | 0.1 | -0.09 | -0.04 | 0.08 | 0 | -0.22 | 0.05 | -0.21 | -0.12 | 0.22 | 0.09 |
| **Reliable_Digit_Span** | -0.07 | 0.02 | -0.11 | 0.1 | -0.09 | -0.25 | -0.23 | 1 | 0.45 | 0.88 | 0.57 | 0.36 | 0.47 | 0.36 | 0.35 | 0.48 | 0.58 | -0.11 | 0.04 |
| **topf_fsiq_z** | -0.11 | 0.07 | 0 | -0.08 | -0.06 | -0.05 | 0.1 | 0.45 | 1 | 0.46 | 0.52 | 0.75 | 0.67 | 0.24 | 0.33 | 0.35 | 0.51 | -0.01 | 0.06 |
| **ds_z** | -0.05 | -0.05 | -0.16 | 0.1 | -0.07 | -0.09 | -0.09 | 0.88 | 0.46 | 1 | 0.57 | 0.42 | 0.39 | 0.4 | 0.43 | 0.54 | 0.57 | -0.12 | 0.01 |
| **ar_z** | -0.19 | 0.02 | -0.28 | -0.29 | 0.14 | -0.05 | -0.04 | 0.57 | 0.52 | 0.57 | 1 | 0.64 | 0.65 | 0.38 | 0.4 | 0.56 | 0.66 | -0.15 | 0.11 |
| **vc_z** | -0.11 | 0.11 | -0.14 | -0.24 | -0.01 | 0.25 | 0.08 | 0.36 | 0.75 | 0.42 | 0.64 | 1 | 0.75 | 0.26 | 0.4 | 0.55 | 0.72 | -0.11 | 0.17 |
| **in_z** | -0.15 | -0.02 | -0.14 | -0.09 | 0.13 | 0.02 | 0 | 0.47 | 0.67 | 0.39 | 0.65 | 0.75 | 1 | 0.24 | 0.5 | 0.47 | 0.53 | 0.02 | 0.03 |
| **sdmt_z** | 0.01 | 0.06 | -0.44 | -0.12 | -0.14 | -0.01 | -0.22 | 0.36 | 0.24 | 0.4 | 0.38 | 0.26 | 0.24 | 1 | 0.32 | 0.43 | 0.42 | -0.09 | 0.02 |
| **vf_letter_z** | -0.07 | -0.12 | -0.22 | 0.13 | 0.15 | 0.18 | 0.05 | 0.35 | 0.33 | 0.43 | 0.4 | 0.4 | 0.5 | 0.32 | 1 | 0.46 | 0.45 | 0.45 | -0.01 |
| **vf_category_z** | -0.03 | 0.15 | -0.37 | 0.05 | 0.14 | 0.18 | -0.21 | 0.48 | 0.35 | 0.54 | 0.56 | 0.55 | 0.47 | 0.43 | 0.46 | 1 | 0.68 | -0.55 | -0.37 |
| **vf_switch_tot_z** | 0.01 | 0.11 | -0.34 | -0.14 | 0.2 | 0.17 | -0.12 | 0.58 | 0.51 | 0.57 | 0.66 | 0.72 | 0.53 | 0.42 | 0.45 | 0.68 | 1 | -0.23 | 0.4 |
| **vf_contrast_lc_z** | -0.05 | -0.23 | 0.15 | 0.05 | 0 | -0.01 | 0.22 | -0.11 | -0.01 | -0.12 | -0.15 | -0.11 | 0.02 | -0.09 | 0.45 | -0.55 | -0.23 | 1 | 0.37 |
| **vf_contrast_sc_z** | 0.04 | -0.04 | -0.02 | -0.31 | 0.17 | 0.01 | 0.09 | 0.04 | 0.06 | 0.01 | 0.11 | 0.17 | 0.03 | 0.02 | -0.01 | -0.37 | 0.4 | 0.37 | 1 |

**Table S3**. Detailed results for correlation analysis when outliers are excluded from the data. Presented are p-values for the cohort of people with FDS.

|  | **gad7_total** | **phq9_total** | **mini_total_risk_score** | **tec_total_presence_score** | **des2_average** | **soms2_symptoms_total_1to53** | **ssq_total_score_4weeksCorrected** | **Reliable_Digit_Span** | **topf_fsiq_z** | **ds_z** | **ar_z** | **vc_z** | **in_z** | **sdmt_z** | **vf_letter_z** | **vf_category_z** | **vf_switch_tot_z** | **vf_contrast_lc_z** | **vf_contrast_sc_z** |
| --- | --- | --- | --- | --- | --- | --- | --- | --- | --- | --- | --- | --- | --- | --- | --- | --- | --- | --- | --- |
| **gad7_total** | NA | 0.001 | 0.051 | 0.049 | 0.429 | 0.505 | 0.797 | 0.692 | 0.512 | 0.746 | 0.251 | 0.5 | 0.357 | 0.963 | 0.66 | 0.84 | 0.964 | 0.774 | 0.803 |
| **phq9_total** | 0.001 | NA | 0.076 | 0.627 | 0.114 | 0.623 | 0.98 | 0.919 | 0.678 | 0.75 | 0.885 | 0.497 | 0.883 | 0.723 | 0.483 | 0.355 | 0.508 | 0.156 | 0.82 |
| **mini_total_risk_score** | 0.051 | 0.076 | NA | 0.112 | 0.239 | 0.857 | 0.112 | 0.491 | 0.984 | 0.337 | 0.082 | 0.386 | 0.391 | 0.005 | 0.188 | 0.021 | 0.036 | 0.352 | 0.9 |
| **tec_total_presence_score** | 0.049 | 0.627 | 0.112 | NA | 0.812 | 0.379 | 0.26 | 0.56 | 0.608 | 0.56 | 0.074 | 0.148 | 0.579 | 0.462 | 0.441 | 0.75 | 0.386 | 0.778 | 0.055 |
| **des2_average** | 0.429 | 0.114 | 0.239 | 0.812 | NA | 0.002 | 0.385 | 0.603 | 0.732 | 0.689 | 0.383 | 0.935 | 0.436 | 0.4 | 0.358 | 0.409 | 0.234 | 0.995 | 0.298 |
| **soms2_symptoms_total_1to53** | 0.505 | 0.623 | 0.857 | 0.379 | 0.002 | NA | 0.011 | 0.128 | 0.787 | 0.579 | 0.747 | 0.136 | 0.887 | 0.965 | 0.27 | 0.277 | 0.312 | 0.935 | 0.937 |
| **ssq_total_score_4weeksCorrected** | 0.797 | 0.98 | 0.112 | 0.26 | 0.385 | 0.011 | NA | 0.153 | 0.532 | 0.576 | 0.818 | 0.65 | 0.981 | 0.192 | 0.783 | 0.192 | 0.461 | 0.183 | 0.58 |
| **Reliable_Digit_Span** | 0.692 | 0.919 | 0.491 | 0.56 | 0.603 | 0.128 | 0.153 | NA | 0.004 | 0 | 0 | 0.027 | 0.002 | 0.024 | 0.028 | 0.002 | 0 | 0.511 | 0.789 |
| **topf_fsiq_z** | 0.512 | 0.678 | 0.984 | 0.608 | 0.732 | 0.787 | 0.532 | 0.004 | NA | 0.004 | 0.001 | 0 | 0 | 0.146 | 0.039 | 0.027 | 0.001 | 0.966 | 0.711 |
| **ds_z** | 0.746 | 0.75 | 0.337 | 0.56 | 0.689 | 0.579 | 0.576 | 0 | 0.004 | NA | 0 | 0.009 | 0.016 | 0.014 | 0.007 | 0 | 0 | 0.49 | 0.948 |
| **ar_z** | 0.251 | 0.885 | 0.082 | 0.074 | 0.383 | 0.747 | 0.818 | 0 | 0.001 | 0 | NA | 0 | 0 | 0.018 | 0.013 | 0 | 0 | 0.371 | 0.486 |
| **vc_z** | 0.5 | 0.497 | 0.386 | 0.148 | 0.935 | 0.136 | 0.65 | 0.027 | 0 | 0.009 | 0 | NA | 0 | 0.12 | 0.012 | 0 | 0 | 0.495 | 0.304 |
| **in_z** | 0.357 | 0.883 | 0.391 | 0.579 | 0.436 | 0.887 | 0.981 | 0.002 | 0 | 0.016 | 0 | 0 | NA | 0.155 | 0.001 | 0.003 | 0.001 | 0.899 | 0.872 |
| **sdmt_z** | 0.963 | 0.723 | 0.005 | 0.462 | 0.4 | 0.965 | 0.192 | 0.024 | 0.146 | 0.014 | 0.018 | 0.12 | 0.155 | NA | 0.053 | 0.008 | 0.009 | 0.582 | 0.916 |
| **vf_letter_z** | 0.66 | 0.483 | 0.188 | 0.441 | 0.358 | 0.27 | 0.783 | 0.028 | 0.039 | 0.007 | 0.013 | 0.012 | 0.001 | 0.053 | NA | 0.003 | 0.004 | 0.004 | 0.939 |
| **vf_category_z** | 0.84 | 0.355 | 0.021 | 0.75 | 0.409 | 0.277 | 0.192 | 0.002 | 0.027 | 0 | 0 | 0 | 0.003 | 0.008 | 0.003 | NA | 0 | 0 | 0.021 |
| **vf_switch_tot_z** | 0.964 | 0.508 | 0.036 | 0.386 | 0.234 | 0.312 | 0.461 | 0 | 0.001 | 0 | 0 | 0 | 0.001 | 0.009 | 0.004 | 0 | NA | 0.171 | 0.013 |
| **vf_contrast_lc_z** | 0.774 | 0.156 | 0.352 | 0.778 | 0.995 | 0.935 | 0.183 | 0.511 | 0.966 | 0.49 | 0.371 | 0.495 | 0.899 | 0.582 | 0.004 | 0 | 0.171 | NA | 0.021 |
| **vf_contrast_sc_z** | 0.803 | 0.82 | 0.9 | 0.055 | 0.298 | 0.937 | 0.58 | 0.789 | 0.711 | 0.948 | 0.486 | 0.304 | 0.872 | 0.916 | 0.939 | 0.021 | 0.013 | 0.021 | NA |

**Table S4**. Detailed results for correlation analysis when outliers are excluded from the data. Presented are the Spearman Correlation Coefficients for the cohort of people with epilepsy

|  | **gad7_total** | **phq9_total** | **mini_total_risk_score** | **tec_total_presence_score** | **des2_average** | **soms2_symptoms_total_1to53** | **ssq_total_score_4weeksCorrected** | **Reliable_Digit_Span** | **topf_fsiq_z** | **ds_z** | **ar_z** | **vc_z** | **in_z** | **sdmt_z** | **vf_letter_z** | **vf_category_z** | **vf_switch_tot_z** | **vf_contrast_lc_z** | **vf_contrast_sc_z** |
| --- | --- | --- | --- | --- | --- | --- | --- | --- | --- | --- | --- | --- | --- | --- | --- | --- | --- | --- | --- |
| **gad7_total** | NA | 0.001 | 0.051 | 0.049 | 0.429 | 0.505 | 0.797 | 0.692 | 0.512 | 0.746 | 0.251 | 0.5 | 0.357 | 0.963 | 0.66 | 0.84 | 0.964 | 0.774 | 0.803 |
| **phq9_total** | 0.001 | NA | 0.076 | 0.627 | 0.114 | 0.623 | 0.98 | 0.919 | 0.678 | 0.75 | 0.885 | 0.497 | 0.883 | 0.723 | 0.483 | 0.355 | 0.508 | 0.156 | 0.82 |
| **mini_total_risk_score** | 0.051 | 0.076 | NA | 0.112 | 0.239 | 0.857 | 0.112 | 0.491 | 0.984 | 0.337 | 0.082 | 0.386 | 0.391 | 0.005 | 0.188 | 0.021 | 0.036 | 0.352 | 0.9 |
| **tec_total_presence_score** | 0.049 | 0.627 | 0.112 | NA | 0.812 | 0.379 | 0.26 | 0.56 | 0.608 | 0.56 | 0.074 | 0.148 | 0.579 | 0.462 | 0.441 | 0.75 | 0.386 | 0.778 | 0.055 |
| **des2_average** | 0.429 | 0.114 | 0.239 | 0.812 | NA | 0.002 | 0.385 | 0.603 | 0.732 | 0.689 | 0.383 | 0.935 | 0.436 | 0.4 | 0.358 | 0.409 | 0.234 | 0.995 | 0.298 |
| **soms2_symptoms_total_1to53** | 0.505 | 0.623 | 0.857 | 0.379 | 0.002 | NA | 0.011 | 0.128 | 0.787 | 0.579 | 0.747 | 0.136 | 0.887 | 0.965 | 0.27 | 0.277 | 0.312 | 0.935 | 0.937 |
| **ssq_total_score_4weeksCorrected** | 0.797 | 0.98 | 0.112 | 0.26 | 0.385 | 0.011 | NA | 0.153 | 0.532 | 0.576 | 0.818 | 0.65 | 0.981 | 0.192 | 0.783 | 0.192 | 0.461 | 0.183 | 0.58 |
| **Reliable_Digit_Span** | 0.692 | 0.919 | 0.491 | 0.56 | 0.603 | 0.128 | 0.153 | NA | 0.004 | 0 | 0 | 0.027 | 0.002 | 0.024 | 0.028 | 0.002 | 0 | 0.511 | 0.789 |
| **topf_fsiq_z** | 0.512 | 0.678 | 0.984 | 0.608 | 0.732 | 0.787 | 0.532 | 0.004 | NA | 0.004 | 0.001 | 0 | 0 | 0.146 | 0.039 | 0.027 | 0.001 | 0.966 | 0.711 |
| **ds_z** | 0.746 | 0.75 | 0.337 | 0.56 | 0.689 | 0.579 | 0.576 | 0 | 0.004 | NA | 0 | 0.009 | 0.016 | 0.014 | 0.007 | 0 | 0 | 0.49 | 0.948 |
| **ar_z** | 0.251 | 0.885 | 0.082 | 0.074 | 0.383 | 0.747 | 0.818 | 0 | 0.001 | 0 | NA | 0 | 0 | 0.018 | 0.013 | 0 | 0 | 0.371 | 0.486 |
| **vc_z** | 0.5 | 0.497 | 0.386 | 0.148 | 0.935 | 0.136 | 0.65 | 0.027 | 0 | 0.009 | 0 | NA | 0 | 0.12 | 0.012 | 0 | 0 | 0.495 | 0.304 |
| **in_z** | 0.357 | 0.883 | 0.391 | 0.579 | 0.436 | 0.887 | 0.981 | 0.002 | 0 | 0.016 | 0 | 0 | NA | 0.155 | 0.001 | 0.003 | 0.001 | 0.899 | 0.872 |
| **sdmt_z** | 0.963 | 0.723 | 0.005 | 0.462 | 0.4 | 0.965 | 0.192 | 0.024 | 0.146 | 0.014 | 0.018 | 0.12 | 0.155 | NA | 0.053 | 0.008 | 0.009 | 0.582 | 0.916 |
| **vf_letter_z** | 0.66 | 0.483 | 0.188 | 0.441 | 0.358 | 0.27 | 0.783 | 0.028 | 0.039 | 0.007 | 0.013 | 0.012 | 0.001 | 0.053 | NA | 0.003 | 0.004 | 0.004 | 0.939 |
| **vf_category_z** | 0.84 | 0.355 | 0.021 | 0.75 | 0.409 | 0.277 | 0.192 | 0.002 | 0.027 | 0 | 0 | 0 | 0.003 | 0.008 | 0.003 | NA | 0 | 0 | 0.021 |
| **vf_switch_tot_z** | 0.964 | 0.508 | 0.036 | 0.386 | 0.234 | 0.312 | 0.461 | 0 | 0.001 | 0 | 0 | 0 | 0.001 | 0.009 | 0.004 | 0 | NA | 0.171 | 0.013 |
| **vf_contrast_lc_z** | 0.774 | 0.156 | 0.352 | 0.778 | 0.995 | 0.935 | 0.183 | 0.511 | 0.966 | 0.49 | 0.371 | 0.495 | 0.899 | 0.582 | 0.004 | 0 | 0.171 | NA | 0.021 |
| **vf_contrast_sc_z** | 0.803 | 0.82 | 0.9 | 0.055 | 0.298 | 0.937 | 0.58 | 0.789 | 0.711 | 0.948 | 0.486 | 0.304 | 0.872 | 0.916 | 0.939 | 0.021 | 0.013 | 0.021 | NA |

**Table S5**. Detailed results for correlation analysis when outliers are excluded from the data. Presented are the p-values for the cohort of people with epilepsy.

|  | **gad7_total** | **phq9_total** | **mini_total_risk_score** | **tec_total_presence_score** | **des2_average** | **soms2_symptoms_total_1to53** | **ssq_total_score_4weeksCorrected** | **Reliable_Digit_Span** | **topf_fsiq_z** | **ds_z** | **ar_z** | **vc_z** | **in_z** | **sdmt_z** | **vf_letter_z** | **vf_category_z** | **vf_switch_tot_z** | **vf_contrast_lc_z** | **vf_contrast_sc_z** |
| --- | --- | --- | --- | --- | --- | --- | --- | --- | --- | --- | --- | --- | --- | --- | --- | --- | --- | --- | --- |
| **gad7_total** | NA | 0 | 0.082 | 0.2 | 0.009 | 0.797 | 0.024 | 0.203 | 0.408 | 0.363 | 0.557 | 0.354 | 0.898 | 0.907 | 0.335 | 0.251 | 0.235 | 0.53 | 0.904 |
| **phq9_total** | 0 | NA | 0.02 | 0.034 | 0.023 | 0.226 | 0.01 | 0.279 | 0.236 | 0.653 | 0.246 | 0.084 | 0.776 | 0.141 | 0.069 | 0.228 | 0.186 | 0.16 | 0.42 |
| **mini_total_risk_score** | 0.082 | 0.02 | NA | 0.271 | 0.326 | 0.342 | 0.184 | 0.744 | 0.668 | 0.887 | 0.206 | 0.382 | 0.77 | 0.686 | 0.743 | 0.543 | 0.419 | 0.214 | 0.849 |
| **tec_total_presence_score** | 0.2 | 0.034 | 0.271 | NA | 0.001 | 0.35 | 0.219 | 0.444 | 0.416 | 0.443 | 0.91 | 0.413 | 0.746 | 0.679 | 0.647 | 0.42 | 0.644 | 0.884 | 0.031 |
| **des2_average** | 0.009 | 0.023 | 0.326 | 0.001 | NA | 0.396 | 0.688 | 0.159 | 0.685 | 0.543 | 0.234 | 0.66 | 0.95 | 0.529 | 0.561 | 0.299 | 0.318 | 0.729 | 0.792 |
| **soms2_symptoms_total_1to53** | 0.797 | 0.226 | 0.342 | 0.35 | 0.396 | NA | 0.311 | 0.152 | 0.111 | 0.484 | 0.017 | 0.017 | 0.004 | 0.208 | 0.96 | 0.079 | 0.147 | 0.124 | 0.503 |
| **ssq_total_score_4weeksCorrected** | 0.024 | 0.01 | 0.184 | 0.219 | 0.688 | 0.311 | NA | 0.864 | 0.903 | 0.786 | 0.474 | 0.406 | 0.554 | 0.227 | 0.61 | 0.15 | 0.995 | 0.719 | 0.123 |
| **Reliable_Digit_Span** | 0.203 | 0.279 | 0.744 | 0.444 | 0.159 | 0.152 | 0.864 | NA | 0.002 | 0 | 0.003 | 0 | 0.001 | 0.06 | 0.006 | 0.007 | 0.009 | 0.766 | 0.269 |
| **topf_fsiq_z** | 0.408 | 0.236 | 0.668 | 0.416 | 0.685 | 0.111 | 0.903 | 0.002 | NA | 0.001 | 0.001 | 0 | 0 | 0.001 | 0.002 | 0.022 | 0.02 | 0.84 | 0.295 |
| **ds_z** | 0.363 | 0.653 | 0.887 | 0.443 | 0.543 | 0.484 | 0.786 | 0 | 0.001 | NA | 0 | 0 | 0.005 | 0.011 | 0.001 | 0.001 | 0.001 | 0.753 | 0.248 |
| **ar_z** | 0.557 | 0.246 | 0.206 | 0.91 | 0.234 | 0.017 | 0.474 | 0.003 | 0.001 | 0 | NA | 0 | 0.001 | 0 | 0.023 | 0 | 0.001 | 0.136 | 0.165 |
| **vc_z** | 0.354 | 0.084 | 0.382 | 0.413 | 0.66 | 0.017 | 0.406 | 0 | 0 | 0 | 0 | NA | 0 | 0.001 | 0.003 | 0.001 | 0.002 | 0.907 | 0.342 |
| **in_z** | 0.898 | 0.776 | 0.77 | 0.746 | 0.95 | 0.004 | 0.554 | 0.001 | 0 | 0.005 | 0.001 | 0 | NA | 0.191 | 0.003 | 0.002 | 0.053 | 0.962 | 0.092 |
| **sdmt_z** | 0.907 | 0.141 | 0.686 | 0.679 | 0.529 | 0.208 | 0.227 | 0.06 | 0.001 | 0.011 | 0 | 0.001 | 0.191 | NA | 0.02 | 0.009 | 0.111 | 0.598 | 0.071 |
| **vf_letter_z** | 0.335 | 0.069 | 0.743 | 0.647 | 0.561 | 0.96 | 0.61 | 0.006 | 0.002 | 0.001 | 0.023 | 0.003 | 0.003 | 0.02 | NA | 0 | 0.029 | 0.012 | 0.006 |
| **vf_category_z** | 0.251 | 0.228 | 0.543 | 0.42 | 0.299 | 0.079 | 0.15 | 0.007 | 0.022 | 0.001 | 0 | 0.001 | 0.002 | 0.009 | 0 | NA | 0 | 0.174 | 0.002 |
| **vf_switch_tot_z** | 0.235 | 0.186 | 0.419 | 0.644 | 0.318 | 0.147 | 0.995 | 0.009 | 0.02 | 0.001 | 0.001 | 0.002 | 0.053 | 0.111 | 0.029 | 0 | NA | 0.065 | 0.121 |
| **vf_contrast_lc_z** | 0.53 | 0.16 | 0.214 | 0.884 | 0.729 | 0.124 | 0.719 | 0.766 | 0.84 | 0.753 | 0.136 | 0.907 | 0.962 | 0.598 | 0.012 | 0.174 | 0.065 | NA | 0.848 |
| **vf_contrast_sc_z** | 0.904 | 0.42 | 0.849 | 0.031 | 0.792 | 0.503 | 0.123 | 0.269 | 0.295 | 0.248 | 0.165 | 0.342 | 0.092 | 0.071 | 0.006 | 0.002 | 0.121 | 0.848 | NA |

**Table S6**. Detailed results for correlation analysis when outliers are excluded from the data. Presented are the Z statistic for the difference between correlation coefficients of people with epilepsy and people with FDS.

|  | **gad7_total** | **phq9_total** | **mini_total_risk_score** | **tec_total_presence_score** | **des2_average** | **soms2_symptoms_total_1to53** | **ssq_total_score_4weeksCorrected** | **Reliable_Digit_Span** | **topf_fsiq_z** | **ds_z** | **ar_z** | **vc_z** | **in_z** | **sdmt_z** | **vf_letter_z** | **vf_category_z** | **vf_switch_tot_z** | **vf_contrast_lc_z** | **vf_contrast_sc_z** |
| --- | --- | --- | --- | --- | --- | --- | --- | --- | --- | --- | --- | --- | --- | --- | --- | --- | --- | --- | --- |
| **gad7_total** | NA | 1.23 | -0.05 | -0.39 | 1.41 | 0.62 | 1.75 | 1.14 | -0.15 | 0.85 | 0.34 | -0.2 | 0.69 | -0.11 | -0.39 | -0.68 | -0.86 | -0.27 | -0.25 |
| **phq9_total** | 1.23 | NA | 0.52 | 1.21 | 0.67 | 0.55 | 1.88 | 0.7 | -1.11 | 0.53 | -0.92 | -1.68 | -0.11 | -1.28 | -0.84 | -1.47 | -1.37 | -0.11 | 0.72 |
| **mini_total_risk_score** | -0.05 | 0.52 | NA | -0.27 | -0.06 | 0.79 | -0.1 | 0.67 | 0.31 | 0.53 | 0.25 | -0.04 | 0.76 | 1.57 | 1.08 | 1.08 | 0.83 | 0.29 | -0.05 |
| **tec_total_presence_score** | -0.39 | 1.21 | -0.27 | NA | 2.52 | 1.24 | 1.59 | 0.16 | 0.9 | 0.15 | 1.08 | 1.52 | 0.13 | 0.77 | -0.82 | -0.78 | 0.89 | -0.29 | 2.77 |
| **des2_average** | 1.41 | 0.67 | -0.06 | 2.52 | NA | -1.41 | -0.27 | 1.35 | 0.51 | 0.7 | -1.42 | 0.37 | -0.55 | 0.1 | -1.01 | -1.3 | -1.5 | -0.25 | -0.49 |
| **soms2_symptoms_total_1to53** | 0.62 | 0.55 | 0.79 | 1.24 | -1.41 | NA | -0.97 | -0.01 | -0.96 | -0.13 | -1.5 | -2.71 | -2.15 | -0.86 | -0.69 | -1.99 | -1.7 | 1.19 | 0.42 |
| **ssq_total_score_4weeksCorrected** | 1.75 | 1.88 | -0.1 | 1.59 | -0.27 | -0.97 | NA | 1.05 | -0.32 | 0.56 | -0.35 | -0.88 | 0.43 | 0.02 | -0.53 | -0.18 | 0.48 | -0.6 | 0.72 |
| **Reliable_Digit_Span** | 1.14 | 0.7 | 0.67 | 0.16 | 1.35 | -0.01 | 1.05 | NA | 0.36 | -0.51 | -0.39 | 1.36 | 0.37 | -0.17 | 0.52 | -0.11 | -0.72 | 0.21 | -0.94 |
| **topf_fsiq_z** | -0.15 | -1.11 | 0.31 | 0.9 | 0.51 | -0.96 | -0.32 | 0.36 | NA | 0.35 | 0.15 | 0.49 | 0.28 | 1.33 | 0.88 | 0.19 | -0.57 | 0.17 | -0.97 |
| **ds_z** | 0.85 | 0.53 | 0.53 | 0.15 | 0.7 | -0.13 | 0.56 | -0.51 | 0.35 | NA | 0.07 | 0.83 | 0.39 | 0.16 | 0.66 | 0.05 | -0.23 | 0.23 | -0.85 |
| **ar_z** | 0.34 | -0.92 | 0.25 | 1.08 | -1.42 | -1.5 | -0.35 | -0.39 | 0.15 | 0.07 | NA | 0.2 | -0.64 | 0.95 | -0.04 | 0.58 | -0.64 | -0.5 | -1.42 |
| **vc_z** | -0.2 | -1.68 | -0.04 | 1.52 | 0.37 | -2.71 | -0.88 | 1.36 | 0.49 | 0.83 | 0.2 | NA | -0.55 | 1.24 | 0.48 | 0.01 | -1.37 | 0.36 | -1.34 |
| **in_z** | 0.69 | -0.11 | 0.76 | 0.13 | -0.55 | -2.15 | 0.43 | 0.37 | 0.28 | 0.39 | -0.64 | -0.55 | NA | -0.02 | -0.04 | 0.24 | -0.96 | -0.05 | -1.29 |
| **sdmt_z** | -0.11 | -1.28 | 1.57 | 0.77 | 0.1 | -0.86 | 0.02 | -0.17 | 1.33 | 0.16 | 0.95 | 1.24 | -0.02 | NA | 0.35 | 0.11 | -0.65 | -0.02 | -1.33 |
| **vf_letter_z** | -0.39 | -0.84 | 1.08 | -0.82 | -1.01 | -0.69 | -0.53 | 0.52 | 0.88 | 0.66 | -0.04 | 0.48 | -0.04 | 0.35 | NA | 1.27 | -0.38 | -0.08 | -1.9 |
| **vf_category_z** | -0.68 | -1.47 | 1.08 | -0.78 | -1.3 | -1.99 | -0.18 | -0.11 | 0.19 | 0.05 | 0.58 | 0.01 | 0.24 | 0.11 | 1.27 | NA | -0.58 | 1.44 | -0.72 |
| **vf_switch_tot_z** | -0.86 | -1.37 | 0.83 | 0.89 | -1.5 | -1.7 | 0.48 | -0.72 | -0.57 | -0.23 | -0.64 | -1.37 | -0.96 | -0.65 | -0.38 | -0.58 | NA | -0.44 | -0.58 |
| **vf_contrast_lc_z** | -0.27 | -0.11 | 0.29 | -0.29 | -0.25 | 1.19 | -0.6 | 0.21 | 0.17 | 0.23 | -0.5 | 0.36 | -0.05 | -0.02 | -0.08 | 1.44 | -0.44 | NA | -1.65 |
| **vf_contrast_sc_z** | -0.25 | 0.72 | -0.05 | 2.77 | -0.49 | 0.42 | 0.72 | -0.94 | -0.97 | -0.85 | -1.42 | -1.34 | -1.29 | -1.33 | -1.9 | -0.72 | -0.58 | -1.65 | NA |

**Table S7**. Detailed results for correlation analysis when outliers are excluded from the data. Presented are the p-values for the difference between correlation coefficients of people with epilepsy and people with FDS.

|  | **gad7_total** | **phq9_total** | **mini_total_risk_score** | **tec_total_presence_score** | **des2_average** | **soms2_symptoms_total_1to53** | **ssq_total_score_4weeksCorrected** | **Reliable_Digit_Span** | **topf_fsiq_z** | **ds_z** | **ar_z** | **vc_z** | **in_z** | **sdmt_z** | **vf_letter_z** | **vf_category_z** | **vf_switch_tot_z** | **vf_contrast_lc_z** | **vf_contrast_sc_z** |
| --- | --- | --- | --- | --- | --- | --- | --- | --- | --- | --- | --- | --- | --- | --- | --- | --- | --- | --- | --- |
| **gad7_total** | NA | 0.218 | 0.959 | 0.696 | 0.158 | 0.535 | 0.079 | 0.252 | 0.879 | 0.397 | 0.737 | 0.839 | 0.493 | 0.912 | 0.699 | 0.494 | 0.391 | 0.79 | 0.806 |
| **phq9_total** | 0.218 | NA | 0.606 | 0.227 | 0.506 | 0.582 | 0.06 | 0.483 | 0.267 | 0.598 | 0.359 | 0.094 | 0.915 | 0.201 | 0.4 | 0.142 | 0.17 | 0.909 | 0.473 |
| **mini_total_risk_score** | 0.959 | 0.606 | NA | 0.79 | 0.955 | 0.428 | 0.919 | 0.502 | 0.756 | 0.595 | 0.804 | 0.967 | 0.448 | 0.115 | 0.279 | 0.28 | 0.409 | 0.769 | 0.959 |
| **tec_total_presence_score** | 0.696 | 0.227 | 0.79 | NA | 0.012 | 0.215 | 0.112 | 0.876 | 0.369 | 0.879 | 0.278 | 0.128 | 0.894 | 0.441 | 0.414 | 0.438 | 0.373 | 0.774 | 0.006 |
| **des2_average** | 0.158 | 0.506 | 0.955 | 0.012 | NA | 0.158 | 0.785 | 0.176 | 0.607 | 0.484 | 0.154 | 0.712 | 0.583 | 0.921 | 0.311 | 0.195 | 0.133 | 0.8 | 0.627 |
| **soms2_symptoms_total_1to53** | 0.535 | 0.582 | 0.428 | 0.215 | 0.158 | NA | 0.333 | 0.989 | 0.338 | 0.9 | 0.134 | 0.007 | 0.031 | 0.387 | 0.491 | 0.047 | 0.089 | 0.235 | 0.673 |
| **ssq_total_score_4weeksCorrected** | 0.079 | 0.06 | 0.919 | 0.112 | 0.785 | 0.333 | NA | 0.296 | 0.749 | 0.579 | 0.726 | 0.381 | 0.669 | 0.988 | 0.594 | 0.859 | 0.631 | 0.546 | 0.471 |
| **Reliable_Digit_Span** | 0.252 | 0.483 | 0.502 | 0.876 | 0.176 | 0.989 | 0.296 | NA | 0.719 | 0.611 | 0.693 | 0.175 | 0.714 | 0.863 | 0.6 | 0.909 | 0.47 | 0.832 | 0.345 |
| **topf_fsiq_z** | 0.879 | 0.267 | 0.756 | 0.369 | 0.607 | 0.338 | 0.749 | 0.719 | NA | 0.726 | 0.879 | 0.621 | 0.782 | 0.185 | 0.379 | 0.847 | 0.566 | 0.863 | 0.332 |
| **ds_z** | 0.397 | 0.598 | 0.595 | 0.879 | 0.484 | 0.9 | 0.579 | 0.611 | 0.726 | NA | 0.941 | 0.407 | 0.695 | 0.874 | 0.51 | 0.959 | 0.819 | 0.821 | 0.396 |
| **ar_z** | 0.737 | 0.359 | 0.804 | 0.278 | 0.154 | 0.134 | 0.726 | 0.693 | 0.879 | 0.941 | NA | 0.841 | 0.52 | 0.34 | 0.972 | 0.559 | 0.525 | 0.617 | 0.155 |
| **vc_z** | 0.839 | 0.094 | 0.967 | 0.128 | 0.712 | 0.007 | 0.381 | 0.175 | 0.621 | 0.407 | 0.841 | NA | 0.585 | 0.214 | 0.635 | 0.994 | 0.171 | 0.716 | 0.181 |
| **in_z** | 0.493 | 0.915 | 0.448 | 0.894 | 0.583 | 0.031 | 0.669 | 0.714 | 0.782 | 0.695 | 0.52 | 0.585 | NA | 0.981 | 0.967 | 0.807 | 0.337 | 0.962 | 0.198 |
| **sdmt_z** | 0.912 | 0.201 | 0.115 | 0.441 | 0.921 | 0.387 | 0.988 | 0.863 | 0.185 | 0.874 | 0.34 | 0.214 | 0.981 | NA | 0.723 | 0.911 | 0.514 | 0.985 | 0.182 |
| **vf_letter_z** | 0.699 | 0.4 | 0.279 | 0.414 | 0.311 | 0.491 | 0.594 | 0.6 | 0.379 | 0.51 | 0.972 | 0.635 | 0.967 | 0.723 | NA | 0.204 | 0.705 | 0.933 | 0.058 |
| **vf_category_z** | 0.494 | 0.142 | 0.28 | 0.438 | 0.195 | 0.047 | 0.859 | 0.909 | 0.847 | 0.959 | 0.559 | 0.994 | 0.807 | 0.911 | 0.204 | NA | 0.562 | 0.15 | 0.47 |
| **vf_switch_tot_z** | 0.391 | 0.17 | 0.409 | 0.373 | 0.133 | 0.089 | 0.631 | 0.47 | 0.566 | 0.819 | 0.525 | 0.171 | 0.337 | 0.514 | 0.705 | 0.562 | NA | 0.662 | 0.565 |
| **vf_contrast_lc_z** | 0.79 | 0.909 | 0.769 | 0.774 | 0.8 | 0.235 | 0.546 | 0.832 | 0.863 | 0.821 | 0.617 | 0.716 | 0.962 | 0.985 | 0.933 | 0.15 | 0.662 | NA | 0.099 |
| **vf_contrast_sc_z** | 0.806 | 0.473 | 0.959 | 0.006 | 0.627 | 0.673 | 0.471 | 0.345 | 0.332 | 0.396 | 0.155 | 0.181 | 0.198 | 0.182 | 0.058 | 0.47 | 0.565 | 0.099 | NA |

**Figure S1.** Graphical representation of Fisher’s z test results when outliers are maintained in the data. Correlation matrices for people with epilepsy (left), people with FDS (middle) and for the difference in correlation coefficients between epilepsy and FDS (right), with associated significance levels (asterisks).

Correlations of interest are those within the black frame. Notes: * Significant at α = 0.05; ** Significant at α = 0.01; *** Significant at α = 0.001. Blue = positive association; Red = negative association; the colour intensity reflects the magnitude of the correlation coefficient (or the magnitude of the difference for the right matrix)

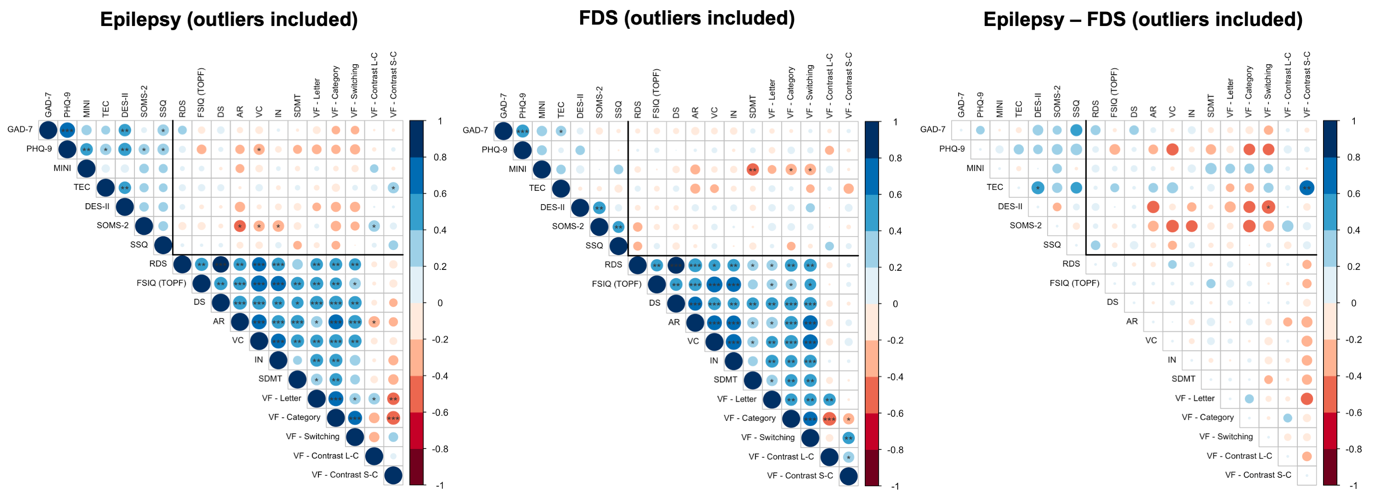

**Table S8**. Detailed results for correlation analysis when outliers are maintained in the data. Presented are the Spearman Correlation Coefficients for the cohort of people with FDS.

|  | **gad7_total** | **phq9_total** | **mini_total_risk_score** | **tec_total_presence_score** | **des2_average** | **soms2_symptoms_total_1to53** | **ssq_total_score_4weeksCorrected** | **Reliable_Digit_Span** | **topf_fsiq_z** | **ds_z** | **ar_z** | **vc_z** | **in_z** | **sdmt_z** | **vf_letter_z** | **vf_category_z** | **vf_switch_tot_z** | **vf_contrast_lc_z** | **vf_contrast_sc_z** |
| --- | --- | --- | --- | --- | --- | --- | --- | --- | --- | --- | --- | --- | --- | --- | --- | --- | --- | --- | --- |
| **gad7_total** | 1 | 0.51 | 0.31 | 0.32 | 0.13 | -0.15 | -0.04 | -0.07 | -0.11 | -0.1 | -0.19 | -0.12 | -0.15 | 0.07 | -0.07 | -0.03 | 0.04 | -0.05 | 0.04 |
| **phq9_total** | 0.51 | 1 | 0.29 | 0.08 | 0.26 | 0 | 0 | 0.02 | 0.07 | 0 | 0.02 | 0.08 | -0.02 | 0.09 | -0.12 | 0.15 | 0.1 | -0.23 | -0.04 |
| **mini_total_risk_score** | 0.31 | 0.29 | 1 | 0.26 | 0.19 | 0 | 0.26 | -0.11 | 0 | -0.15 | -0.28 | -0.11 | -0.14 | -0.45 | -0.22 | -0.37 | -0.39 | 0.15 | -0.02 |
| **tec_total_presence_score** | 0.32 | 0.08 | 0.26 | 1 | -0.04 | -0.07 | -0.18 | 0.1 | -0.08 | 0.11 | -0.29 | -0.22 | -0.09 | -0.09 | 0.13 | 0.05 | -0.21 | 0.05 | -0.31 |
| **des2_average** | 0.13 | 0.26 | 0.19 | -0.04 | 1 | 0.47 | 0.14 | -0.09 | -0.06 | -0.07 | 0.14 | 0.06 | 0.13 | -0.11 | 0.15 | 0.14 | 0.24 | 0 | 0.17 |
| **soms2_symptoms_total_1to53** | -0.15 | 0 | 0 | -0.07 | 0.47 | 1 | 0.42 | -0.26 | -0.1 | -0.11 | -0.08 | 0.11 | 0.01 | -0.01 | 0.18 | 0.16 | 0.12 | 0 | -0.05 |
| **ssq_total_score_4weeksCorrected** | -0.04 | 0 | 0.26 | -0.18 | 0.14 | 0.42 | 1 | -0.23 | 0.1 | -0.15 | -0.04 | 0.09 | 0 | -0.14 | 0.05 | -0.21 | -0.06 | 0.22 | 0.09 |
| **Reliable_Digit_Span** | -0.07 | 0.02 | -0.11 | 0.1 | -0.09 | -0.26 | -0.23 | 1 | 0.45 | 0.88 | 0.57 | 0.41 | 0.47 | 0.36 | 0.35 | 0.48 | 0.5 | -0.11 | 0.04 |
| **topf_fsiq_z** | -0.11 | 0.07 | 0 | -0.08 | -0.06 | -0.1 | 0.1 | 0.45 | 1 | 0.49 | 0.52 | 0.77 | 0.67 | 0.29 | 0.33 | 0.35 | 0.41 | -0.01 | 0.06 |
| **ds_z** | -0.1 | 0 | -0.15 | 0.11 | -0.07 | -0.11 | -0.15 | 0.88 | 0.49 | 1 | 0.6 | 0.51 | 0.44 | 0.46 | 0.47 | 0.57 | 0.51 | -0.12 | -0.04 |
| **ar_z** | -0.19 | 0.02 | -0.28 | -0.29 | 0.14 | -0.08 | -0.04 | 0.57 | 0.52 | 0.6 | 1 | 0.67 | 0.65 | 0.37 | 0.4 | 0.56 | 0.68 | -0.15 | 0.11 |
| **vc_z** | -0.12 | 0.08 | -0.11 | -0.22 | 0.06 | 0.11 | 0.09 | 0.41 | 0.77 | 0.51 | 0.67 | 1 | 0.76 | 0.36 | 0.43 | 0.58 | 0.71 | -0.14 | 0.19 |
| **in_z** | -0.15 | -0.02 | -0.14 | -0.09 | 0.13 | 0.01 | 0 | 0.47 | 0.67 | 0.44 | 0.65 | 0.76 | 1 | 0.29 | 0.5 | 0.47 | 0.51 | 0.02 | 0.03 |
| **sdmt_z** | 0.07 | 0.09 | -0.45 | -0.09 | -0.11 | -0.01 | -0.14 | 0.36 | 0.29 | 0.46 | 0.37 | 0.36 | 0.29 | 1 | 0.37 | 0.47 | 0.49 | -0.13 | -0.01 |
| **vf_letter_z** | -0.07 | -0.12 | -0.22 | 0.13 | 0.15 | 0.18 | 0.05 | 0.35 | 0.33 | 0.47 | 0.4 | 0.43 | 0.5 | 0.37 | 1 | 0.46 | 0.47 | 0.45 | -0.01 |
| **vf_category_z** | -0.03 | 0.15 | -0.37 | 0.05 | 0.14 | 0.16 | -0.21 | 0.48 | 0.35 | 0.57 | 0.56 | 0.58 | 0.47 | 0.47 | 0.46 | 1 | 0.63 | -0.55 | -0.37 |
| **vf_switch_tot_z** | 0.04 | 0.1 | -0.39 | -0.21 | 0.24 | 0.12 | -0.06 | 0.5 | 0.41 | 0.51 | 0.68 | 0.71 | 0.51 | 0.49 | 0.47 | 0.63 | 1 | -0.16 | 0.45 |
| **vf_contrast_lc_z** | -0.05 | -0.23 | 0.15 | 0.05 | 0 | 0 | 0.22 | -0.11 | -0.01 | -0.12 | -0.15 | -0.14 | 0.02 | -0.13 | 0.45 | -0.55 | -0.16 | 1 | 0.37 |
| **vf_contrast_sc_z** | 0.04 | -0.04 | -0.02 | -0.31 | 0.17 | -0.05 | 0.09 | 0.04 | 0.06 | -0.04 | 0.11 | 0.19 | 0.03 | -0.01 | -0.01 | -0.37 | 0.45 | 0.37 | 1 |

**Table S9**. Detailed results for correlation analysis when outliers are maintained in the data. Presented are the p-values for the cohort of people with FDS.

|  | **gad7_total** | **phq9_total** | **mini_total_risk_score** | **tec_total_presence_score** | **des2_average** | **soms2_symptoms_total_1to53** | **ssq_total_score_4weeksCorrected** | **Reliable_Digit_Span** | **topf_fsiq_z** | **ds_z** | **ar_z** | **vc_z** | **in_z** | **sdmt_z** | **vf_letter_z** | **vf_category_z** | **vf_switch_tot_z** | **vf_contrast_lc_z** | **vf_contrast_sc_z** |
| --- | --- | --- | --- | --- | --- | --- | --- | --- | --- | --- | --- | --- | --- | --- | --- | --- | --- | --- | --- |
| **gad7_total** | NA | 0.001 | 0.051 | 0.049 | 0.429 | 0.378 | 0.797 | 0.692 | 0.512 | 0.545 | 0.251 | 0.45 | 0.357 | 0.672 | 0.66 | 0.84 | 0.808 | 0.774 | 0.803 |
| **phq9_total** | 0.001 | NA | 0.076 | 0.627 | 0.114 | 0.997 | 0.98 | 0.919 | 0.678 | 0.996 | 0.885 | 0.612 | 0.883 | 0.603 | 0.483 | 0.355 | 0.526 | 0.156 | 0.82 |
| **mini_total_risk_score** | 0.051 | 0.076 | NA | 0.112 | 0.239 | 0.996 | 0.112 | 0.491 | 0.984 | 0.367 | 0.082 | 0.502 | 0.391 | 0.004 | 0.188 | 0.021 | 0.015 | 0.352 | 0.9 |
| **tec_total_presence_score** | 0.049 | 0.627 | 0.112 | NA | 0.812 | 0.691 | 0.26 | 0.56 | 0.608 | 0.516 | 0.074 | 0.188 | 0.579 | 0.592 | 0.441 | 0.75 | 0.207 | 0.778 | 0.055 |
| **des2_average** | 0.429 | 0.114 | 0.239 | 0.812 | NA | 0.003 | 0.385 | 0.603 | 0.732 | 0.662 | 0.383 | 0.704 | 0.436 | 0.518 | 0.358 | 0.409 | 0.146 | 0.995 | 0.298 |
| **soms2_symptoms_total_1to53** | 0.378 | 0.997 | 0.996 | 0.691 | 0.003 | NA | 0.008 | 0.114 | 0.554 | 0.496 | 0.633 | 0.49 | 0.943 | 0.949 | 0.271 | 0.331 | 0.459 | 0.998 | 0.755 |
| **ssq_total_score_4weeksCorrected** | 0.797 | 0.98 | 0.112 | 0.26 | 0.385 | 0.008 | NA | 0.153 | 0.532 | 0.357 | 0.818 | 0.594 | 0.981 | 0.38 | 0.783 | 0.192 | 0.726 | 0.183 | 0.58 |
| **Reliable_Digit_Span** | 0.692 | 0.919 | 0.491 | 0.56 | 0.603 | 0.114 | 0.153 | NA | 0.004 | 0 | 0 | 0.01 | 0.002 | 0.024 | 0.028 | 0.002 | 0.001 | 0.511 | 0.789 |
| **topf_fsiq_z** | 0.512 | 0.678 | 0.984 | 0.608 | 0.732 | 0.554 | 0.532 | 0.004 | NA | 0.002 | 0.001 | 0 | 0 | 0.077 | 0.039 | 0.027 | 0.01 | 0.966 | 0.711 |
| **ds_z** | 0.545 | 0.996 | 0.367 | 0.516 | 0.662 | 0.496 | 0.357 | 0 | 0.002 | NA | 0 | 0.001 | 0.005 | 0.004 | 0.002 | 0 | 0.001 | 0.479 | 0.83 |
| **ar_z** | 0.251 | 0.885 | 0.082 | 0.074 | 0.383 | 0.633 | 0.818 | 0 | 0.001 | 0 | NA | 0 | 0 | 0.019 | 0.013 | 0 | 0 | 0.371 | 0.486 |
| **vc_z** | 0.45 | 0.612 | 0.502 | 0.188 | 0.704 | 0.49 | 0.594 | 0.01 | 0 | 0.001 | 0 | NA | 0 | 0.026 | 0.006 | 0 | 0 | 0.385 | 0.258 |
| **in_z** | 0.357 | 0.883 | 0.391 | 0.579 | 0.436 | 0.943 | 0.981 | 0.002 | 0 | 0.005 | 0 | 0 | NA | 0.078 | 0.001 | 0.003 | 0.001 | 0.899 | 0.872 |
| **sdmt_z** | 0.672 | 0.603 | 0.004 | 0.592 | 0.518 | 0.949 | 0.38 | 0.024 | 0.077 | 0.004 | 0.019 | 0.026 | 0.078 | NA | 0.021 | 0.003 | 0.002 | 0.427 | 0.947 |
| **vf_letter_z** | 0.66 | 0.483 | 0.188 | 0.441 | 0.358 | 0.271 | 0.783 | 0.028 | 0.039 | 0.002 | 0.013 | 0.006 | 0.001 | 0.021 | NA | 0.003 | 0.002 | 0.004 | 0.939 |
| **vf_category_z** | 0.84 | 0.355 | 0.021 | 0.75 | 0.409 | 0.331 | 0.192 | 0.002 | 0.027 | 0 | 0 | 0 | 0.003 | 0.003 | 0.003 | NA | 0 | 0 | 0.021 |
| **vf_switch_tot_z** | 0.808 | 0.526 | 0.015 | 0.207 | 0.146 | 0.459 | 0.726 | 0.001 | 0.01 | 0.001 | 0 | 0 | 0.001 | 0.002 | 0.002 | 0 | NA | 0.325 | 0.004 |
| **vf_contrast_lc_z** | 0.774 | 0.156 | 0.352 | 0.778 | 0.995 | 0.998 | 0.183 | 0.511 | 0.966 | 0.479 | 0.371 | 0.385 | 0.899 | 0.427 | 0.004 | 0 | 0.325 | NA | 0.021 |
| **vf_contrast_sc_z** | 0.803 | 0.82 | 0.9 | 0.055 | 0.298 | 0.755 | 0.58 | 0.789 | 0.711 | 0.83 | 0.486 | 0.258 | 0.872 | 0.947 | 0.939 | 0.021 | 0.004 | 0.021 | NA |

**Table S10**. Detailed results for correlation analysis when outliers are maintained in the data. Presented are the Spearman Correlation Coefficients for the cohort of people with epilepsy.

|  | **gad7_total** | **phq9_total** | **mini_total_risk_score** | **tec_total_presence_score** | **des2_average** | **soms2_symptoms_total_1to53** | **ssq_total_score_4weeksCorrected** | **Reliable_Digit_Span** | **topf_fsiq_z** | **ds_z** | **ar_z** | **vc_z** | **in_z** | **sdmt_z** | **vf_letter_z** | **vf_category_z** | **vf_switch_tot_z** | **vf_contrast_lc_z** | **vf_contrast_sc_z** |
| --- | --- | --- | --- | --- | --- | --- | --- | --- | --- | --- | --- | --- | --- | --- | --- | --- | --- | --- | --- |
| **gad7_total** | 1 | 0.72 | 0.3 | 0.23 | 0.46 | 0.09 | 0.39 | 0.22 | -0.15 | 0.16 | -0.1 | -0.16 | 0.02 | -0.02 | -0.17 | -0.2 | -0.21 | -0.01 | -0.02 |
| **phq9_total** | 0.72 | 1 | 0.47 | 0.38 | 0.52 | 0.34 | 0.39 | 0.13 | -0.26 | 0.06 | -0.25 | -0.35 | -0.12 | -0.23 | -0.27 | -0.25 | -0.3 | -0.04 | 0.05 |
| **mini_total_risk_score** | 0.3 | 0.47 | 1 | 0.19 | 0.2 | 0.26 | 0.23 | 0.06 | 0.08 | -0.03 | -0.22 | -0.15 | 0.05 | -0.07 | 0.06 | -0.08 | -0.14 | 0.2 | -0.03 |
| **tec_total_presence_score** | 0.23 | 0.38 | 0.19 | 1 | 0.46 | 0.24 | 0.22 | 0.14 | 0.14 | 0.14 | -0.02 | 0.15 | -0.06 | 0.07 | -0.08 | -0.19 | 0.08 | 0.12 | 0.37 |
| **des2_average** | 0.46 | 0.52 | 0.2 | 0.46 | 1 | 0.26 | 0.27 | 0.08 | -0.05 | -0.03 | -0.31 | -0.06 | -0.12 | -0.16 | -0.21 | -0.3 | -0.31 | 0.01 | 0.02 |
| **soms2_symptoms_total_1to53** | 0.09 | 0.34 | 0.26 | 0.24 | 0.26 | 1 | 0.28 | -0.18 | -0.18 | -0.06 | -0.42 | -0.36 | -0.39 | -0.16 | 0.06 | -0.31 | -0.24 | 0.38 | 0.14 |
| **ssq_total_score_4weeksCorrected** | 0.39 | 0.39 | 0.23 | 0.22 | 0.27 | 0.28 | 1 | 0.03 | 0.02 | 0.05 | -0.13 | -0.15 | 0.11 | -0.21 | -0.09 | -0.22 | 0 | 0.06 | 0.27 |
| **Reliable_Digit_Span** | 0.22 | 0.13 | 0.06 | 0.14 | 0.08 | -0.18 | 0.03 | 1 | 0.52 | 0.84 | 0.5 | 0.62 | 0.54 | 0.33 | 0.46 | 0.48 | 0.44 | -0.09 | -0.2 |
| **topf_fsiq_z** | -0.15 | -0.26 | 0.08 | 0.14 | -0.05 | -0.18 | 0.02 | 0.52 | 1 | 0.53 | 0.54 | 0.8 | 0.71 | 0.53 | 0.52 | 0.44 | 0.4 | -0.03 | -0.18 |
| **ds_z** | 0.16 | 0.06 | -0.03 | 0.14 | -0.03 | -0.06 | 0.05 | 0.84 | 0.53 | 1 | 0.58 | 0.58 | 0.47 | 0.43 | 0.56 | 0.58 | 0.53 | -0.09 | -0.2 |
| **ar_z** | -0.1 | -0.25 | -0.22 | -0.02 | -0.31 | -0.42 | -0.13 | 0.5 | 0.54 | 0.58 | 1 | 0.67 | 0.55 | 0.57 | 0.39 | 0.68 | 0.56 | -0.38 | -0.24 |
| **vc_z** | -0.16 | -0.35 | -0.15 | 0.15 | -0.06 | -0.36 | -0.15 | 0.62 | 0.8 | 0.58 | 0.67 | 1 | 0.68 | 0.53 | 0.5 | 0.57 | 0.51 | -0.12 | -0.17 |
| **in_z** | 0.02 | -0.12 | 0.05 | -0.06 | -0.12 | -0.39 | 0.11 | 0.54 | 0.71 | 0.47 | 0.55 | 0.68 | 1 | 0.23 | 0.49 | 0.51 | 0.33 | -0.03 | -0.29 |
| **sdmt_z** | -0.02 | -0.23 | -0.07 | 0.07 | -0.16 | -0.16 | -0.21 | 0.33 | 0.53 | 0.43 | 0.57 | 0.53 | 0.23 | 1 | 0.4 | 0.48 | 0.28 | -0.16 | -0.31 |
| **vf_letter_z** | -0.17 | -0.27 | 0.06 | -0.08 | -0.21 | 0.06 | -0.09 | 0.46 | 0.52 | 0.56 | 0.39 | 0.5 | 0.49 | 0.4 | 1 | 0.69 | 0.37 | 0.39 | -0.46 |
| **vf_category_z** | -0.2 | -0.25 | -0.08 | -0.19 | -0.3 | -0.31 | -0.22 | 0.48 | 0.44 | 0.58 | 0.68 | 0.57 | 0.51 | 0.48 | 0.69 | 1 | 0.6 | -0.33 | -0.56 |
| **vf_switch_tot_z** | -0.21 | -0.3 | -0.14 | 0.08 | -0.31 | -0.24 | 0 | 0.44 | 0.4 | 0.53 | 0.56 | 0.51 | 0.33 | 0.28 | 0.37 | 0.6 | 1 | -0.31 | 0.27 |
| **vf_contrast_lc_z** | -0.01 | -0.04 | 0.2 | 0.12 | 0.01 | 0.38 | 0.06 | -0.09 | -0.03 | -0.09 | -0.38 | -0.12 | -0.03 | -0.16 | 0.39 | -0.33 | -0.31 | 1 | 0.1 |
| **vf_contrast_sc_z** | -0.02 | 0.05 | -0.03 | 0.37 | 0.02 | 0.14 | 0.27 | -0.2 | -0.18 | -0.2 | -0.24 | -0.17 | -0.29 | -0.31 | -0.46 | -0.56 | 0.27 | 0.1 | 1 |

**Table S11**. Detailed results for correlation analysis when outliers are maintained in the data. Presented are the p-values for the cohort of people with epilepsy.

|  | **gad7_total** | **phq9_total** | **mini_total_risk_score** | **tec_total_presence_score** | **des2_average** | **soms2_symptoms_total_1to53** | **ssq_total_score_4weeksCorrected** | **Reliable_Digit_Span** | **topf_fsiq_z** | **ds_z** | **ar_z** | **vc_z** | **in_z** | **sdmt_z** | **vf_letter_z** | **vf_category_z** | **vf_switch_tot_z** | **vf_contrast_lc_z** | **vf_contrast_sc_z** |
| --- | --- | --- | --- | --- | --- | --- | --- | --- | --- | --- | --- | --- | --- | --- | --- | --- | --- | --- | --- |
| **gad7_total** | NA | 0 | 0.082 | 0.2 | 0.006 | 0.623 | 0.024 | 0.203 | 0.408 | 0.363 | 0.557 | 0.354 | 0.898 | 0.907 | 0.335 | 0.245 | 0.235 | 0.959 | 0.904 |
| **phq9_total** | 0 | NA | 0.005 | 0.028 | 0.002 | 0.047 | 0.021 | 0.449 | 0.132 | 0.728 | 0.158 | 0.041 | 0.506 | 0.192 | 0.12 | 0.157 | 0.082 | 0.808 | 0.797 |
| **mini_total_risk_score** | 0.082 | 0.005 | NA | 0.271 | 0.268 | 0.134 | 0.184 | 0.744 | 0.668 | 0.887 | 0.206 | 0.382 | 0.77 | 0.686 | 0.743 | 0.663 | 0.419 | 0.25 | 0.849 |
| **tec_total_presence_score** | 0.2 | 0.028 | 0.271 | NA | 0.006 | 0.17 | 0.219 | 0.444 | 0.416 | 0.443 | 0.91 | 0.413 | 0.746 | 0.679 | 0.647 | 0.282 | 0.644 | 0.501 | 0.031 |
| **des2_average** | 0.006 | 0.002 | 0.268 | 0.006 | NA | 0.144 | 0.124 | 0.645 | 0.766 | 0.879 | 0.075 | 0.733 | 0.484 | 0.362 | 0.237 | 0.089 | 0.076 | 0.968 | 0.925 |
| **soms2_symptoms_total_1to53** | 0.623 | 0.047 | 0.134 | 0.17 | 0.144 | NA | 0.115 | 0.303 | 0.316 | 0.75 | 0.014 | 0.039 | 0.022 | 0.353 | 0.742 | 0.072 | 0.178 | 0.026 | 0.437 |
| **ssq_total_score_4weeksCorrected** | 0.024 | 0.021 | 0.184 | 0.219 | 0.124 | 0.115 | NA | 0.864 | 0.903 | 0.786 | 0.474 | 0.406 | 0.554 | 0.227 | 0.61 | 0.221 | 0.995 | 0.734 | 0.123 |
| **Reliable_Digit_Span** | 0.203 | 0.449 | 0.744 | 0.444 | 0.645 | 0.303 | 0.864 | NA | 0.002 | 0 | 0.003 | 0 | 0.001 | 0.06 | 0.006 | 0.004 | 0.009 | 0.615 | 0.269 |
| **topf_fsiq_z** | 0.408 | 0.132 | 0.668 | 0.416 | 0.766 | 0.316 | 0.903 | 0.002 | NA | 0.001 | 0.001 | 0 | 0 | 0.001 | 0.002 | 0.01 | 0.02 | 0.846 | 0.295 |
| **ds_z** | 0.363 | 0.728 | 0.887 | 0.443 | 0.879 | 0.75 | 0.786 | 0 | 0.001 | NA | 0 | 0 | 0.005 | 0.011 | 0.001 | 0 | 0.001 | 0.628 | 0.248 |
| **ar_z** | 0.557 | 0.158 | 0.206 | 0.91 | 0.075 | 0.014 | 0.474 | 0.003 | 0.001 | 0 | NA | 0 | 0.001 | 0 | 0.023 | 0 | 0.001 | 0.027 | 0.165 |
| **vc_z** | 0.354 | 0.041 | 0.382 | 0.413 | 0.733 | 0.039 | 0.406 | 0 | 0 | 0 | 0 | NA | 0 | 0.001 | 0.003 | 0 | 0.002 | 0.482 | 0.342 |
| **in_z** | 0.898 | 0.506 | 0.77 | 0.746 | 0.484 | 0.022 | 0.554 | 0.001 | 0 | 0.005 | 0.001 | 0 | NA | 0.191 | 0.003 | 0.002 | 0.053 | 0.856 | 0.092 |
| **sdmt_z** | 0.907 | 0.192 | 0.686 | 0.679 | 0.362 | 0.353 | 0.227 | 0.06 | 0.001 | 0.011 | 0 | 0.001 | 0.191 | NA | 0.02 | 0.004 | 0.111 | 0.375 | 0.071 |
| **vf_letter_z** | 0.335 | 0.12 | 0.743 | 0.647 | 0.237 | 0.742 | 0.61 | 0.006 | 0.002 | 0.001 | 0.023 | 0.003 | 0.003 | 0.02 | NA | 0 | 0.029 | 0.022 | 0.006 |
| **vf_category_z** | 0.245 | 0.157 | 0.663 | 0.282 | 0.089 | 0.072 | 0.221 | 0.004 | 0.01 | 0 | 0 | 0 | 0.002 | 0.004 | 0 | NA | 0 | 0.056 | 0.001 |
| **vf_switch_tot_z** | 0.235 | 0.082 | 0.419 | 0.644 | 0.076 | 0.178 | 0.995 | 0.009 | 0.02 | 0.001 | 0.001 | 0.002 | 0.053 | 0.111 | 0.029 | 0 | NA | 0.073 | 0.121 |
| **vf_contrast_lc_z** | 0.959 | 0.808 | 0.25 | 0.501 | 0.968 | 0.026 | 0.734 | 0.615 | 0.846 | 0.628 | 0.027 | 0.482 | 0.856 | 0.375 | 0.022 | 0.056 | 0.073 | NA | 0.569 |
| **vf_contrast_sc_z** | 0.904 | 0.797 | 0.849 | 0.031 | 0.925 | 0.437 | 0.123 | 0.269 | 0.295 | 0.248 | 0.165 | 0.342 | 0.092 | 0.071 | 0.006 | 0.001 | 0.121 | 0.569 | NA |

**Table S12**. Detailed results for correlation analysis when outliers are maintained in the data. Presented are the Z statistic for the difference between correlation coefficients of people with epilepsy and people with FDS.

|  | **gad7_total** | **phq9_total** | **mini_total_risk_score** | **tec_total_presence_score** | **des2_average** | **soms2_symptoms_total_1to53** | **ssq_total_score_4weeksCorrected** | **Reliable_Digit_Span** | **topf_fsiq_z** | **ds_z** | **ar_z** | **vc_z** | **in_z** | **sdmt_z** | **vf_letter_z** | **vf_category_z** | **vf_switch_tot_z** | **vf_contrast_lc_z** | **vf_contrast_sc_z** |
| --- | --- | --- | --- | --- | --- | --- | --- | --- | --- | --- | --- | --- | --- | --- | --- | --- | --- | --- | --- |
| **gad7_total** | NA | 1.34 | -0.05 | -0.39 | 1.45 | 0.91 | 1.75 | 1.14 | -0.15 | 1.03 | 0.34 | -0.16 | 0.69 | -0.35 | -0.39 | -0.68 | -0.99 | 0.15 | -0.25 |
| **phq9_total** | 1.34 | NA | 0.84 | 1.23 | 1.21 | 1.39 | 1.64 | 0.46 | -1.32 | 0.24 | -1.08 | -1.77 | -0.37 | -1.25 | -0.63 | -1.59 | -1.63 | 0.75 | 0.33 |
| **mini_total_risk_score** | -0.05 | 0.84 | NA | -0.27 | 0.01 | 1.05 | -0.1 | 0.67 | 0.31 | 0.48 | 0.25 | -0.17 | 0.76 | 1.62 | 1.08 | 1.21 | 1.03 | 0.2 | -0.05 |
| **tec_total_presence_score** | -0.39 | 1.23 | -0.27 | NA | 2.11 | 1.22 | 1.59 | 0.16 | 0.9 | 0.11 | 1.08 | 1.42 | 0.13 | 0.63 | -0.82 | -0.96 | 1.14 | 0.29 | 2.77 |
| **des2_average** | 1.45 | 1.21 | 0.01 | 2.11 | NA | -0.97 | 0.52 | 0.66 | 0.01 | 0.18 | -1.81 | -0.48 | -0.99 | -0.22 | -1.42 | -1.72 | -2.19 | 0.03 | -0.61 |
| **soms2_symptoms_total_1to53** | 0.91 | 1.39 | 1.05 | 1.22 | -0.97 | NA | -0.64 | 0.31 | -0.32 | 0.22 | -1.43 | -1.9 | -1.67 | -0.61 | -0.48 | -1.89 | -1.42 | 1.58 | 0.74 |
| **ssq_total_score_4weeksCorrected** | 1.75 | 1.64 | -0.1 | 1.59 | 0.52 | -0.64 | NA | 1.05 | -0.32 | 0.78 | -0.35 | -0.92 | 0.43 | -0.28 | -0.53 | -0.01 | 0.22 | -0.63 | 0.72 |
| **Reliable_Digit_Span** | 1.14 | 0.46 | 0.67 | 0.16 | 0.66 | 0.31 | 1.05 | NA | 0.36 | -0.67 | -0.39 | 1.13 | 0.37 | -0.15 | 0.52 | -0.01 | -0.3 | 0.07 | -0.94 |
| **topf_fsiq_z** | -0.15 | -1.32 | 0.31 | 0.9 | 0.01 | -0.32 | -0.32 | 0.36 | NA | 0.21 | 0.15 | 0.32 | 0.28 | 1.13 | 0.88 | 0.38 | -0.04 | -0.11 | -0.97 |
| **ds_z** | 1.03 | 0.24 | 0.48 | 0.11 | 0.18 | 0.22 | 0.78 | -0.67 | 0.21 | NA | -0.12 | 0.42 | 0.17 | -0.11 | 0.44 | 0.03 | 0.06 | 0.12 | -0.67 |
| **ar_z** | 0.34 | -1.08 | 0.25 | 1.08 | -1.81 | -1.43 | -0.35 | -0.39 | 0.15 | -0.12 | NA | 0.02 | -0.64 | 0.99 | -0.04 | 0.8 | -0.79 | -0.98 | -1.42 |
| **vc_z** | -0.16 | -1.77 | -0.17 | 1.42 | -0.48 | -1.9 | -0.92 | 1.13 | 0.32 | 0.42 | 0.02 | NA | -0.72 | 0.82 | 0.35 | -0.08 | -1.23 | 0.07 | -1.4 |
| **in_z** | 0.69 | -0.37 | 0.76 | 0.13 | -0.99 | -1.67 | 0.43 | 0.37 | 0.28 | 0.17 | -0.64 | -0.72 | NA | -0.23 | -0.04 | 0.23 | -0.83 | -0.21 | -1.29 |
| **sdmt_z** | -0.35 | -1.25 | 1.62 | 0.63 | -0.22 | -0.61 | -0.28 | -0.15 | 1.13 | -0.11 | 0.99 | 0.82 | -0.23 | NA | 0.13 | 0.06 | -0.96 | -0.1 | -1.22 |
| **vf_letter_z** | -0.39 | -0.63 | 1.08 | -0.82 | -1.42 | -0.48 | -0.53 | 0.52 | 0.88 | 0.44 | -0.04 | 0.35 | -0.04 | 0.13 | NA | 1.37 | -0.47 | -0.3 | -1.9 |
| **vf_category_z** | -0.68 | -1.59 | 1.21 | -0.96 | -1.72 | -1.89 | -0.01 | -0.01 | 0.38 | 0.03 | 0.8 | -0.08 | 0.23 | 0.06 | 1.37 | NA | -0.19 | 1.08 | -0.95 |
| **vf_switch_tot_z** | -0.99 | -1.63 | 1.03 | 1.14 | -2.19 | -1.42 | 0.22 | -0.3 | -0.04 | 0.06 | -0.79 | -1.23 | -0.83 | -0.96 | -0.47 | -0.19 | NA | -0.62 | -0.79 |
| **vf_contrast_lc_z** | 0.15 | 0.75 | 0.2 | 0.29 | 0.03 | 1.58 | -0.63 | 0.07 | -0.11 | 0.12 | -0.98 | 0.07 | -0.21 | -0.1 | -0.3 | 1.08 | -0.62 | NA | -1.12 |
| **vf_contrast_sc_z** | -0.25 | 0.33 | -0.05 | 2.77 | -0.61 | 0.74 | 0.72 | -0.94 | -0.97 | -0.67 | -1.42 | -1.4 | -1.29 | -1.22 | -1.9 | -0.95 | -0.79 | -1.12 | NA |

**Table S13**. Detailed results for correlation analysis when outliers are maintained in the data. Presented are the p-values for the difference between correlation coefficients of people with epilepsy and people with FDS.

|  | **gad7_total** | **phq9_total** | **mini_total_risk_score** | **tec_total_presence_score** | **des2_average** | **soms2_symptoms_total_1to53** | **ssq_total_score_4weeksCorrected** | **Reliable_Digit_Span** | **topf_fsiq_z** | **ds_z** | **ar_z** | **vc_z** | **in_z** | **sdmt_z** | **vf_letter_z** | **vf_category_z** | **vf_switch_tot_z** | **vf_contrast_lc_z** | **vf_contrast_sc_z** |
| --- | --- | --- | --- | --- | --- | --- | --- | --- | --- | --- | --- | --- | --- | --- | --- | --- | --- | --- | --- |
| **gad7_total** | NA | 0.182 | 0.959 | 0.696 | 0.148 | 0.361 | 0.079 | 0.252 | 0.879 | 0.305 | 0.737 | 0.875 | 0.493 | 0.723 | 0.699 | 0.496 | 0.324 | 0.881 | 0.806 |
| **phq9_total** | 0.182 | NA | 0.401 | 0.217 | 0.226 | 0.164 | 0.101 | 0.644 | 0.186 | 0.811 | 0.279 | 0.077 | 0.713 | 0.212 | 0.526 | 0.112 | 0.104 | 0.453 | 0.744 |
| **mini_total_risk_score** | 0.959 | 0.401 | NA | 0.79 | 0.993 | 0.296 | 0.919 | 0.502 | 0.756 | 0.628 | 0.804 | 0.861 | 0.448 | 0.105 | 0.279 | 0.227 | 0.302 | 0.841 | 0.959 |
| **tec_total_presence_score** | 0.696 | 0.217 | 0.79 | NA | 0.035 | 0.224 | 0.112 | 0.876 | 0.369 | 0.909 | 0.278 | 0.154 | 0.894 | 0.526 | 0.414 | 0.339 | 0.254 | 0.774 | 0.006 |
| **des2_average** | 0.148 | 0.226 | 0.993 | 0.035 | NA | 0.333 | 0.606 | 0.511 | 0.988 | 0.86 | 0.07 | 0.629 | 0.322 | 0.828 | 0.155 | 0.085 | 0.029 | 0.974 | 0.543 |
| **soms2_symptoms_total_1to53** | 0.361 | 0.164 | 0.296 | 0.224 | 0.333 | NA | 0.523 | 0.759 | 0.752 | 0.827 | 0.152 | 0.058 | 0.096 | 0.544 | 0.628 | 0.058 | 0.156 | 0.115 | 0.458 |
| **ssq_total_score_4weeksCorrected** | 0.079 | 0.101 | 0.919 | 0.112 | 0.606 | 0.523 | NA | 0.296 | 0.749 | 0.433 | 0.726 | 0.356 | 0.669 | 0.783 | 0.594 | 0.993 | 0.824 | 0.531 | 0.471 |
| **Reliable_Digit_Span** | 0.252 | 0.644 | 0.502 | 0.876 | 0.511 | 0.759 | 0.296 | NA | 0.719 | 0.502 | 0.693 | 0.258 | 0.714 | 0.877 | 0.6 | 0.99 | 0.764 | 0.94 | 0.345 |
| **topf_fsiq_z** | 0.879 | 0.186 | 0.756 | 0.369 | 0.988 | 0.752 | 0.749 | 0.719 | NA | 0.833 | 0.879 | 0.749 | 0.782 | 0.257 | 0.379 | 0.704 | 0.966 | 0.914 | 0.332 |
| **ds_z** | 0.305 | 0.811 | 0.628 | 0.909 | 0.86 | 0.827 | 0.433 | 0.502 | 0.833 | NA | 0.903 | 0.677 | 0.863 | 0.912 | 0.658 | 0.974 | 0.952 | 0.904 | 0.504 |
| **ar_z** | 0.737 | 0.279 | 0.804 | 0.278 | 0.07 | 0.152 | 0.726 | 0.693 | 0.879 | 0.903 | NA | 0.986 | 0.52 | 0.323 | 0.972 | 0.425 | 0.431 | 0.325 | 0.155 |
| **vc_z** | 0.875 | 0.077 | 0.861 | 0.154 | 0.629 | 0.058 | 0.356 | 0.258 | 0.749 | 0.677 | 0.986 | NA | 0.471 | 0.41 | 0.729 | 0.936 | 0.22 | 0.942 | 0.163 |
| **in_z** | 0.493 | 0.713 | 0.448 | 0.894 | 0.322 | 0.096 | 0.669 | 0.714 | 0.782 | 0.863 | 0.52 | 0.471 | NA | 0.816 | 0.967 | 0.82 | 0.404 | 0.835 | 0.198 |
| **sdmt_z** | 0.723 | 0.212 | 0.105 | 0.526 | 0.828 | 0.544 | 0.783 | 0.877 | 0.257 | 0.912 | 0.323 | 0.41 | 0.816 | NA | 0.9 | 0.951 | 0.335 | 0.917 | 0.222 |
| **vf_letter_z** | 0.699 | 0.526 | 0.279 | 0.414 | 0.155 | 0.628 | 0.594 | 0.6 | 0.379 | 0.658 | 0.972 | 0.729 | 0.967 | 0.9 | NA | 0.171 | 0.639 | 0.761 | 0.058 |
| **vf_category_z** | 0.496 | 0.112 | 0.227 | 0.339 | 0.085 | 0.058 | 0.993 | 0.99 | 0.704 | 0.974 | 0.425 | 0.936 | 0.82 | 0.951 | 0.171 | NA | 0.852 | 0.279 | 0.344 |
| **vf_switch_tot_z** | 0.324 | 0.104 | 0.302 | 0.254 | 0.029 | 0.156 | 0.824 | 0.764 | 0.966 | 0.952 | 0.431 | 0.22 | 0.404 | 0.335 | 0.639 | 0.852 | NA | 0.534 | 0.428 |
| **vf_contrast_lc_z** | 0.881 | 0.453 | 0.841 | 0.774 | 0.974 | 0.115 | 0.531 | 0.94 | 0.914 | 0.904 | 0.325 | 0.942 | 0.835 | 0.917 | 0.761 | 0.279 | 0.534 | NA | 0.264 |
| **vf_contrast_sc_z** | 0.806 | 0.744 | 0.959 | 0.006 | 0.543 | 0.458 | 0.471 | 0.345 | 0.332 | 0.504 | 0.155 | 0.163 | 0.198 | 0.222 | 0.058 | 0.344 | 0.428 | 0.264 | NA |
